# Central Adiposity And Infective Endocarditis: A Cohort Study of UK Biobank Participants

**DOI:** 10.64898/2026.04.22.26351534

**Authors:** Wang Song, Jinwei Zhang, Wei ZhiPeng, Peng Sun, Zhang Ke, Xu Chenzhen, Yue Chuanjie, Yuzhe Zhang, Lingyao Li, Liaoming He, Jianbo Yu, Yongqiang Lai, Hao Cui, Changwei Ren

**Affiliations:** Department of Cardiovascular Surgery Center, Capital Medical University Affiliated Beijing Anzhen Hospital, Beijing Institute of Heart, Lung and Blood Vascular Diseases, Beijing, China

**Keywords:** Infective Endocarditis, Central adiposity, Obesity, Waist circumference, UK Biobank

## Abstract

**Aims:** While traditional anthropometric indices are established cardiovascular predictors, their prognostic value for incident infective endocarditis (IE) remains undefined.

**Methods:** We included 386,859 participants (mean age 57.0 years; 52.9% female) from the UK Biobank between 2006 and 2010 with standardized baseline data on BMI, waist circumference (WC), waist-to-height ratio (WhtR), and the triglyceride-glucose (TyG) index.Multivariable Cox proportional hazard models with restricted cubic splines were used to estimate the hazard ratio (HR) of these indices, adjusting for demographic and clinical risk factors.

**Results:** Over 16.87 median years (25^th^, 16.02; 75^th^, 17.60 percentile) of follow-up, there were a total of 1,124 incident IE events. During the follow-up period, 38,342 total deaths were recorded, of which 8,524 were cardiovascular disease (CVD)-related.Overall, compared to individuals with normal weight and baseline metabolic indices, those in the fourth quartile of WC, WHtR, and TyG index exhibited the highest risk of incident IE. Compared to other metabolic indices, WC (HR = 1.53, 95% CI 1.23 – 1.90,P < 0.001) and WHtR (HR = 1.46, 95% CI 1.20 – 1.78,P < 0.001) demonstrated higher relative increases in risk associated with IE. Furthermore, the risk of IE was significantly elevated among the younger population with abdominal obesity and concomitant diabetes. However, no significant increase in IE risk was observed among participants with pre-existing valvular heart disease (P = 0.796).

**Conclusion:** Compared with BMI, higher WC and WHtR were robustly associated with increased risk of IE, even after adjusting for traditional risk factors. Furthermore, the risk of IE was markedly elevated among younger individuals with abdominal obesity and diabetes.

**Graphical abstract:** 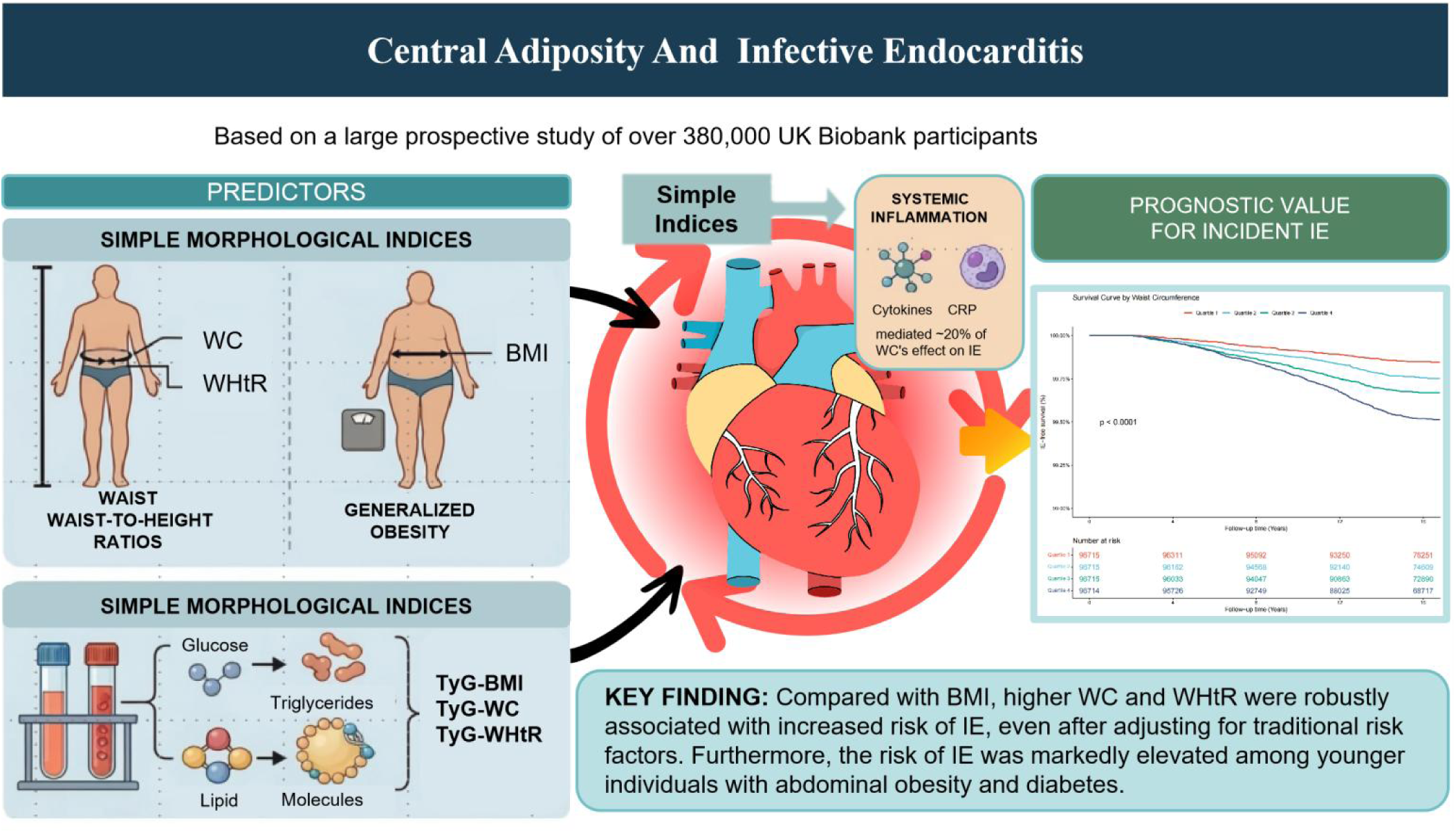

## Introduction

Infective endocarditis (IE) is a devastating cardiovascular infection characterized by substantial morbidity and mortality. Despite advances in diagnostic imaging and antimicrobial therapies, the global annual incidence of IE continues to rise, reaching approximately 13.8 cases per 100,000 individuals, with short-term mortality rates remaining alarmingly high at nearly 30%. Traditionally driven by rheumatic heart disease, the epidemiological landscape of IE has progressively shifted toward older populations burdened with a high prevalence of metabolic comorbidities^1,2^. In light of this evolving profile, identifying novel, modifiable risk factors is imperative for early risk stratification and targeted prevention.

Obesity, conventionally assessed via body mass index (BMI), is a well-established driver of systemic inflammation and cardiovascular disease^3–5^. However, BMI serves as a crude proxy that fails to differentiate between adiposity and lean mass or account for regional fat distribution. While markers of central adiposity, such as waist circumference (WC) and waist-to-height ratio (WHtR), offer superior insights into visceral fat accumulation^6,7^, they do not directly capture the concomitant metabolic dysfunction^8,9^.

Insulin resistance (IR) plays a pivotal role in the pathogenesis of various cardiovascular and infectious diseases^10–13^. Beyond its central role in metabolic syndrome, IR fosters a pro-inflammatory milieu, exacerbates oxidative stress, and impairs endothelial integrity by diminishing nitric oxide bioavailability^14^. Crucially, endothelial injury is the initiating step in the formation of sterile vegetations—the prerequisite nidus for bacterial adherence in IE^15^. Furthermore, IR compromises host immune defenses, thereby potentially increasing susceptibility to the sustained bacteremia necessary for IE development^16,17^.

Recent paradigm shifts in cardiovascular prevention increasingly advocate for the use of complex composite indices—such as the triglyceride-glucose (TyG) index combined with obesity indices (e.g., TyG-WC)—to capture the synergistic deleterious effects of anatomical adiposity and systemic IR^18–22^. While these complex metabolic composites have demonstrated remarkable prognostic value for atherosclerotic cardiovascular diseases^23,24^, it remains largely unexplored whether this “complex is better” paradigm extends to infectious cardiovascular diseases such as IE. Unlike lipid-driven atheroma plaque formation in atherosclerotic cardiovascular diseases, the pathophysiology of IE is primarily driven by endothelial injury and immune dysregulation. Consequently, it is of immense clinical relevance to determine whether calculating complex metabolic formulas provides incremental prognostic value over simple morphological measurements for IE risk stratification.

To address this critical knowledge gap, we leveraged the large-scale, deeply phenotyped UK Biobank prospective cohort to comprehensively evaluate and compare the long-term associations of traditional morphological indices (BMI, WC, WHtR) and complex metabolic composites (TyG-BMI, TyG-WC, TyG-WHtR) with the risk of incident IE. The overall conceptual framework, study design, and primary findings regarding central adiposity and infective endocarditis risk are visually summarized in the Graphical Abstract.

## Methods

### Study population

The UK Biobank is a large-scale, prospective, population-based cohort study designed to investigate the genetic, environmental, and lifestyle determinants of a myriad of diseases. Between 2006 and 2010, over 500,000 community-dwelling adults aged 40–69 years were recruited across 22 assessment centers in England, Wales, and Scotland. At baseline, participants completed comprehensive touch-screen questionnaires and underwent standardized physical examinations—including detailed anthropometric measurements (e.g., height, weight, and waist circumference)—along with the provision of biological samples (blood, urine, and saliva). Longitudinal follow-up was achieved through linkage to national electronic health records and death registries^25^. The study protocol was approved by the North West Multi-centre Research Ethics Committee, and written informed consent was obtained from all participants. Detailed information regarding the study rationale and design is available online (http://www.ukbiobank.ac.uk).

Of the 501,936 initially enrolled participants, we applied a priori sequential exclusion criteria to establish the analytic cohort. Participants were excluded if they: (1) had missing data on core exposure variables, including BMI, waist circumference, height, or components of the TyG index (n = 74,980); or (2) had a baseline diagnosis of cancer (n = 37,671). Furthermore, to minimize potential reverse causality bias, we excluded individuals with less than 2 years of follow-up or those with a documented history of infective endocarditis prior to baseline (n = 2,426). Consequently, the final analytic cohort comprised 386,859 participants(Fig.S6).

### Assessment of TyG and Obesity-Related Indices

The TyG index was calculated using the established formula: ln [fasting triglycerides (mg/dL) × fasting glucose (mg/dL) / 2]^26^. Traditional anthropometric obesity indices included BMI, calculated as weight in kilograms divided by height in meters squared (kg/m²); WC, measured at the midpoint between the lower costal margin and the iliac crest; and WHtR, calculated as WC (cm) divided by height (cm). To systematically integrate metabolic dysfunction with anatomical adiposity, composite TyG-obesity indices—specifically TyG-BMI, TyG-WC, and TyG-WHtR—were generated by multiplying the TyG index by each respective anthropometric parameter^27^. For statistical modeling, all indices were evaluated as continuous variables, standardized and expressed per 1-SD increment. For categorical analyses, BMI was stratified into six clinically established groups: underweight ( <18.5kg/m^2^), normal weight (18.5-24.9 kg/m^2^), overweight (25.0 -29.9 kg/m^2^), and obesity classes I (30.0-34.9kg/m^2^), II (35.0-39.9 kg/m^2^), and III (≥40.0 kg/m^2^)^28^. All remaining indices were divided into quartiles, with the lowest quartile (Q1) serving as the reference group.

### Assessment of Outcomes

The primary outcome was the incidence of IE. Incident IE events were ascertained through linkage to national hospital inpatient records and death registries, utilizing the International Classification of Diseases, 10th Revision (ICD-10) codes I33 (acute and subacute infective endocarditis), I38 (endocarditis, valve unspecified), and I39 (endocarditis and heart valve disorders in diseases classified elsewhere). Person-time at risk was calculated from the date of the baseline assessment until the date of the first IE diagnosis, death from any cause, whichever occurred first.

### Assessment of Covariates

Comprehensive information on potential confounders was collected at baseline using standardized touchscreen questionnaires, nurse-led verbal interviews, and physical examinations.

Sociodemographic and Lifestyle Factors: Sociodemographic variables included age, sex, ethnicity, education level, housing conditions, and the Townsend Deprivation Index (TDI)—a census-based composite measure of area-level socioeconomic deprivation. Lifestyle factors evaluated were smoking status (never, previous, or current), alcohol consumption, and physical activity (quantified via the International Physical Activity Questionnaire). Given the well-established pathogenic link between oral dysbiosis and IE^29,30^, baseline oral health status was rigorously phenotyped using a composite indicator. Participants were classified as having “poor oral health” if they met either of the following criteria prior to baseline: (1) a hospital diagnosis of dental pathology, identified by ICD-10 codes for dental caries (K02), gingivitis and periodontal diseases (K05), other disorders of teeth and supporting structures (K08), or stomatitis (K12); or (2) self-reported oral problems, including bleeding or painful gums, loose teeth, or toothache.

### Clinical and Anthropometric Characteristics

Physical measurements included BMI and WC, alongside resting systolic and diastolic blood pressure obtained via an automated device. Prevalent comorbidities—specifically hypertension, diabetes, and valvular heart disease—were defined using a combination of self-reported medical history and linked hospital admission records (ICD-10 codes). The concurrent use of medications, including antihypertensive agents, lipid-lowering drugs, and antidiabetic medications (e.g., insulin), was also recorded.

### Biochemical Markers

Baseline blood samples were assayed to determine systemic biomarker concentrations, including C-reactive protein (CRP), low-density lipoprotein cholesterol (LDL-C), and high-density lipoprotein cholesterol (HDL-C).

### Statistical Analysis Descriptive Statistics

Continuous variables were summarized as means ± SD or medians with interquartile ranges (IQR), depending on data distribution. Categorical variables were presented as frequencies and percentages. To minimize potential bias and loss of statistical power, missing baseline covariate data were handled using multiple imputation by chained equations, generating five imputed datasets for the main analyses.

### Primary Analysis

Multivariable Cox proportional hazards regression models were utilized to estimate hazard ratios (HRs) and 95% confidence intervals (CIs) for the independent associations between morphological/metabolic obesity indices and the risk of incident IE. The indices were evaluated both as continuous variables (standardized per 1-SD increment) and categorical variables (quartiles). We constructed three progressively adjusted models: Model 1 was unadjusted; Model 2 adjusted for demographic and lifestyle factors (age, sex, ethnicity, education, Townsend deprivation index, smoking status, alcohol consumption, and physical activity); and Model 3 further adjusted for baseline comorbidities (diabetes, hypertension, hypercholesterolemia, valvular heart disease, cardiovascular disease, and poor oral health).No significant multicollinearity was detected among the covariates, with all variance inflation factor values remaining below 5. Kaplan-Meier survival curves were plotted across subgroups and compared using the log-rank test.

### Non-linear Relationships and Phenotype Stratification

To explore potential non-linear dose-response relationships, restricted cubic splines (RCS) with four knots (at the 5th, 35th, 65th, and 95th percentiles) were employed. Subgroup analyses were performed across various clinical strata. Furthermore, participants were categorized into four distinct metabolic-obesity phenotypes: metabolically healthy normal weight (MHNW), metabolically unhealthy normal weight (MUNW), metabolically healthy obesity (MHO), and metabolically unhealthy obesity (MUO)^31^. Clinical obesity was defined as a BMI ≥ 30 kg/m², and metabolic unhealthiness was operationally defined as a TyG index above the cohort median.

### Sensitivity Analyses

To ensure the robustness of our findings, four rigorous sensitivity analyses were conducted: (1) excluding participants with a follow-up duration of less than 2 years to minimize reverse causation; (2) employing a Fine-Gray competing risk regression model to account for the competing risk of all-cause mortality; (3) performing a complete-case analysis (CCA) utilizing only participants without missing data; and (4) calculating E-values to quantify the minimum strength of association that an unmeasured confounder would need to have with both the exposure and the outcome to fully explain away the observed associations.

To explore potential underlying mechanisms, a formal mediation analysis was conducted using a counterfactual framework to quantify the proportion of the association between central adiposity (waist circumference) and incident IE that was mediated by systemic inflammation (log-transformed CRP). We employed 5,000 bootstrap resamples across the full analytical cohort to estimate the average direct effect, average causal mediation effect, and the proportion mediated, along with their 95% CI.

All statistical analyses were performed using R software (version 4.5.1). A two-sided P-value < 0.05 was considered statistically significant.

## Results

### Study Population and Follow-up

The initial UK Biobank cohort comprised 501,936 participants. After applying a priori sequential exclusion criteria—specifically, missing data on core exposures (n = 74,980), a baseline diagnosis of cancer (n = 37,671), and a follow-up duration of less than 2 years to mitigate potential reverse causation (n = 2,426) — the final analytic cohort consisted of 386,859 eligible individuals (Fig.S6). Over a median follow-up of 16.87 years (IQR, 16.02 – 17.60 years), which yielded more than 6.32 million person-years at risk, we identified 1,124 incident cases of IE. The overall crude incidence rate of IE in this cohort was 17.78 per 100,000 person-years.

### Baseline Characteristics

The baseline characteristics of the analytic cohort, stratified by quartiles of WC and WHtR, are presented in Table 1 and Table 2, respectively. The median age of the study population was 57.0 years (IQR, 50.0–63.0 years). Pronounced differences in demographic, lifestyle, and clinical profiles were evident across the quartiles of both central adiposity indices.

**Table 1.** Baseline characteristics of the study individuals according to waist circumference quartiles.

| Variables | Overall | Q1 | Q2 | Q3 | Q4 | P value |
| --- | --- | --- | --- | --- | --- | --- |
| N | 386859 | 96715 | 96715 | 96715 | 96714 |  |
| Age(years) | 57.00 [50.00, 63.00] | 55.00 [48.00, 62.00] | 57.00 [49.00, 63.00] | 58.00 [50.00, 63.00] | 59.00 [51.00, 64.00] | <0.001 |
| **Sex** |  |  |  |  |  |  |
| Female | 204606 (52.9) | 86805 (89.8) | 56701 (58.6) | 34387 (35.6) | 26713 (27.6) | <0.001 |
| Male | 182253 (47.1) | 9910 (10.2) | 40014 (41.4) | 62328 (64.4) | 70001 (72.4) |  |
| **Race** |  |  |  |  |  |  |
| Non-White | 34284 (8.9) | 8401 (8.7) | 9103 (9.4) | 8747 (9.0) | 8033 (8.3) | <0.001 |
| White | 352575 (91.1) | 88314 (91.3) | 87612 (90.6) | 87968 (91.0) | 88681 (91.7) |  |
| TDI | -2.14 [-3.65, 0.53] | -2.35 [-3.76, 0.08] | -2.21 [-3.68, 0.37] | -2.16 [-3.65, 0.50] | -1.78 [-3.45, 1.18] | <0.001 |
| **Education** |  |  |  |  |  |  |
| High | 71770 (18.6) | 14280 (14.8) | 16981 (17.6) | 19523 (20.2) | 20986 (21.7) | <0.001 |
| Low | 144271 (37.3) | 33630 (34.8) | 35463 (36.7) | 36765 (38.0) | 38413 (39.7) |  |
| Medium | 170818 (44.2) | 48805 (50.5) | 44271 (45.8) | 40427 (41.8) | 37315 (38.6) |  |
| **Smoking** |  |  |  |  |  |  |
| Current | 41061 (10.6) | 9157 (9.5) | 10272 (10.6) | 10596 (11.0) | 11036 (11.4) | <0.001 |
| Never | 213203 (55.1) | 60565 (62.6) | 56291 (58.2) | 51322 (53.1) | 45025 (46.6) |  |
| Previous | 132595 (34.3) | 26993 (27.9) | 30152 (31.2) | 34797 (36.0) | 40653 (42.0) |  |
| **Drinking** |  |  |  |  |  |  |
| Current | 356173 (92.1) | 89201 (92.2) | 89343 (92.4) | 89568 (92.6) | 88061 (91.1) | <0.001 |
| Never | 17066 (4.4) | 4464 (4.6) | 4293 (4.4) | 3993 (4.1) | 4316 (4.5) |  |
| Previous | 13620 (3.5) | 3050 (3.2) | 3079 (3.2) | 3154 (3.3) | 4337 (4.5) |  |
| Physical activity<br>(MET-hour/week) | 19.80 [1.65, 48.25] | 23.77 [3.85, 53.00] | 21.90 [2.55, 51.10] | 19.87 [2.20, 48.20] | 14.22 [0.00, 39.65] | <0.001 |
| BMI (kg/m<sup>2</sup>) | 27.44 (4.78) | 23.13 (2.37) | 25.90 (2.59) | 28.05 (2.88) | 32.70 (4.66) | <0.001 |
| SBP(mmHg) | 137.00 [125.00,<br>151.00] | 130.00 [118.00,<br>144.00] | 136.00 [124.00,<br>150.00] | 140.00 [128.00,<br>153.00] | 143.00 [131.00,<br>155.00] | <0.001 |
| DBP(mmHg) | 82.00 [75.00, 89.00] | 77.00 [71.00, 84.00] | 81.00 [74.00, 88.00] | 83.00 [77.00, 90.00] | 86.00 [79.00, 93.00] | <0.001 |
| Log_CRP | 0.35 [-0.29, 1.04] | -0.15 [-0.69, 0.48] | 0.23 [-0.36, 0.88] | 0.45 [-0.13, 1.09] | 0.84 [0.25, 1.48] | <0.001 |
| TC (mmol/L) | 5.65 [4.91, 6.42] | 5.68 [5.00, 6.41] | 5.74 [5.02, 6.50] | 5.69 [4.93, 6.46] | 5.46 [4.66, 6.28] | <0.001 |
| LDL(mmol/L) | 1.75 [1.47, 2.06] | 1.72 [1.46, 2.01] | 1.79 [1.51, 2.09] | 1.79 [1.50, 2.10] | 1.72 [1.41, 2.04] | <0.001 |
| HDL(mmol/L) | 1.40 [1.17, 1.67] | 1.68 [1.44, 1.94] | 1.46 [1.25, 1.70] | 1.31 [1.13, 1.53] | 1.19 [1.03, 1.39] | <0.001 |
| Hba1c (mmol/L) | 35.20 [32.70, 37.90] | 34.30 [32.10, 36.60] | 34.90 [32.50, 37.20] | 35.30 [32.90, 38.00] | 36.60 [33.80, 40.10] | <0.001 |
| Hypertension | 215803 (55.8) | 35908 (37.1) | 49339 (51.0) | 59454 (61.5) | 71102 (73.5) | <0.001 |
| Diabetes | 23474 (6.1) | 1907 (2.0) | 3156 (3.3) | 5404 (5.6) | 13007 (13.4) | <0.001 |
| Hyperlipidemia | 143898 (37.2) | 35315 (36.5) | 39050 (40.4) | 37106 (38.4) | 32427 (33.5) | <0.001 |
| Valve heart disease | 1340 (0.3) | 365 (0.4) | 337 (0.3) | 335 (0.3) | 303 (0.3) | 0.123 |
| Cardiovascular disease | 16639 (4.3) | 1718 (1.8) | 3004 (3.1) | 4514 (4.7) | 7403 (7.7) | <0.001 |
| Oral problems | 81441 (21.1) | 19724 (20.4) | 19829 (20.5) | 19729 (20.4) | 22159 (22.9) | <0.001 |
| Antihypertensive medication | 35392 (9.1) | 7704 (8.0) | 9600 (9.9) | 8450 (8.7) | 9638 (10.0) | <0.001 |
| Insulin | 1607 (0.4) | 368 (0.4) | 285 (0.3) | 308 (0.3) | 646 (0.7) | <0.001 |
| Cholesterol-lowering medication | 25562 (6.6) | 5575 (5.8) | 7012 (7.3) | 6215 (6.4) | 6760 (7.0) | <0.001 |

**Table 2.**
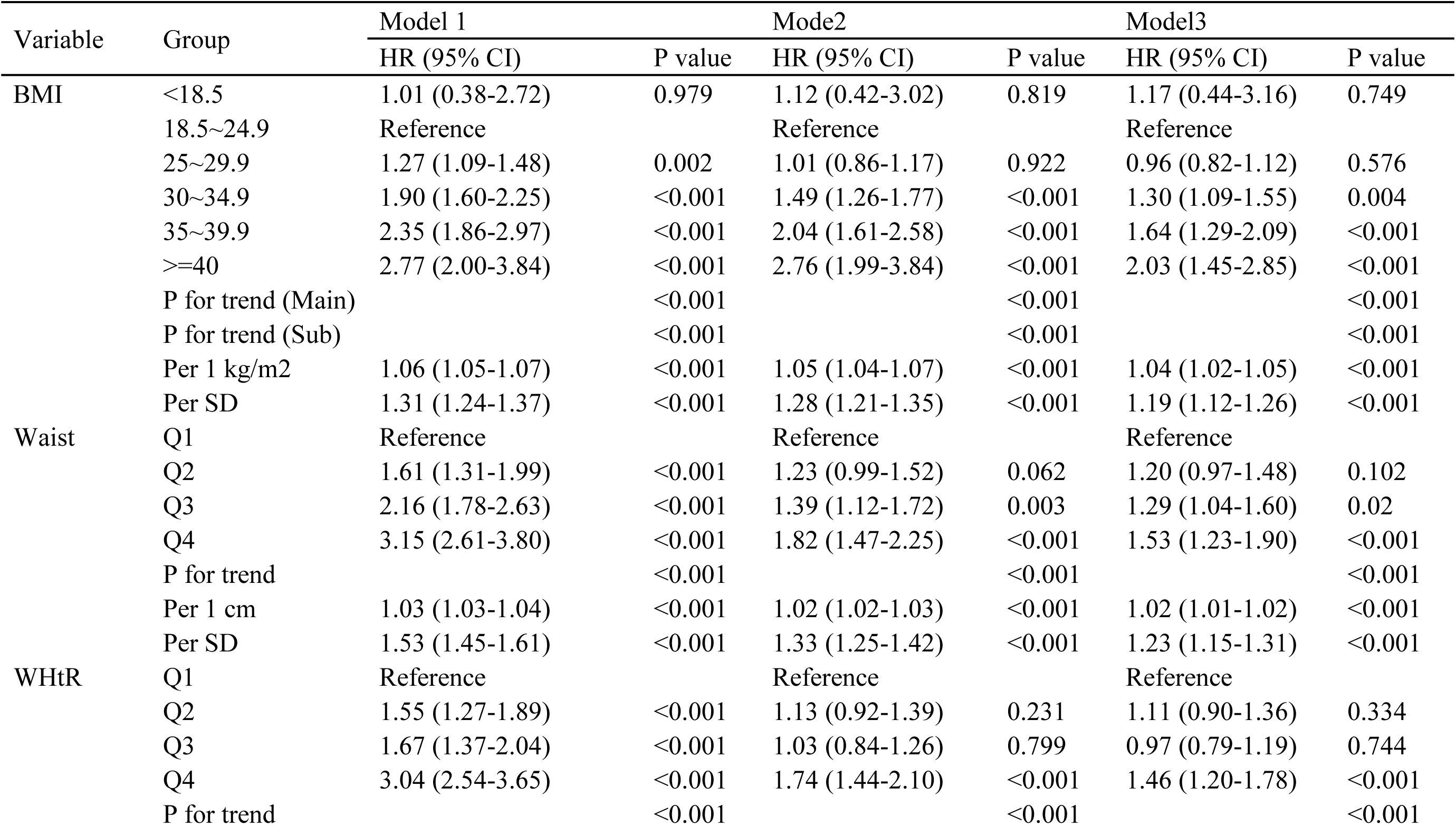
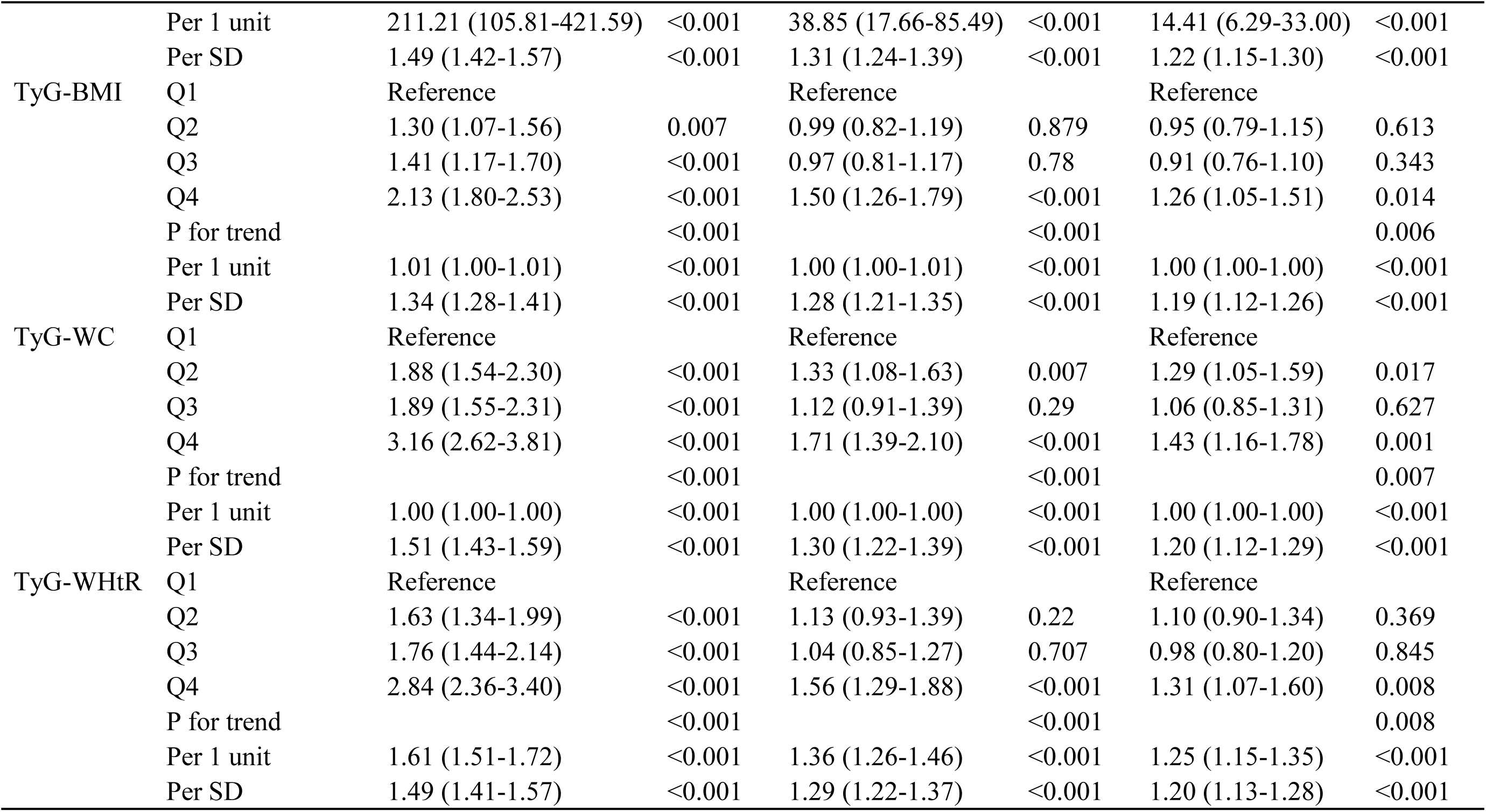
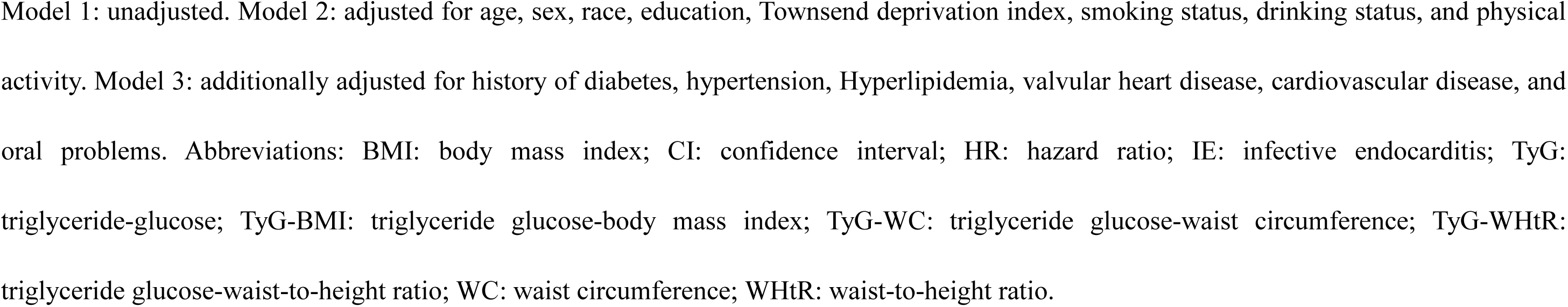
Association between obesity and TyG-related indices and incident infective endocarditis.

Taking WC as a primary example (Table 1), participants in the highest quartile (Q4) were significantly older than those in the lowest quartile (Q1) (median age 59.0 vs. 55.0 years) and comprised a substantially higher proportion of men (72.4% vs. 10.2%). Furthermore, individuals in the upper quartiles exhibited a progressively greater burden of adverse lifestyle factors and cardiometabolic risk markers. Across increasing quartiles of both WC and WHtR, there were significant linear elevations in systolic blood pressure, systemic inflammation (assessed via log-transformed C-reactive protein), and HbA1c levels, coupled with a steady decline in HDL-C (all P < 0.001). Correspondingly, the prevalence of baseline comorbidities was markedly higher in the upper quartiles for both metrics. Most notably, the prevalence of diabetes (13.4% in Q4 vs. 2.0% in Q1 for WC) and cardiovascular disease peaked in the highest quartiles (all P < 0.001). Baseline characteristics stratified by the other morphological and metabolic indices are provided in Supplementary Tables S1–S4, S9.

### Kaplan-Meier Survival Analyses

Kaplan-Meier survival analyses revealed significant differences in the cumulative incidence of IE across the distinct categories of obesity and TyG-related indices (Figure 1). Notably, participants in the highest quartiles of central adiposity metrics—specifically WC (Figure 1B) and WHtR (Figure 1C)—exhibited a markedly higher cumulative risk of incident IE over the follow-up period compared to those in the lowest quartiles (log-rank P < 0.001). Similar graded increases in the cumulative incidence of IE were observed across the quartiles of TyG-WC and TyG-WHtR (log-rank P < 0.001, Figures 1E and 1F).

**Fig. 1.**
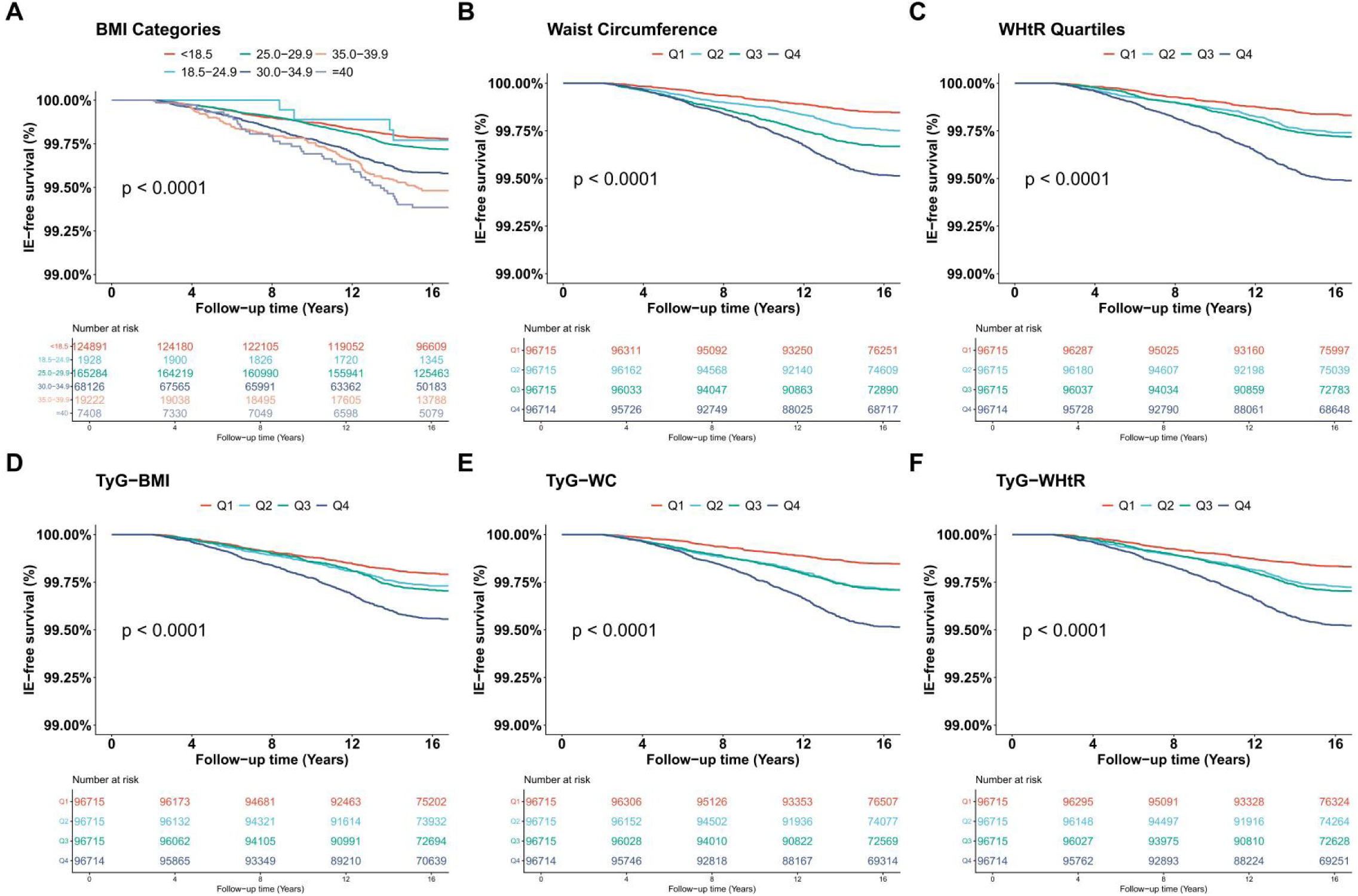
Kaplan-Meier surival curves for incident infective endocarditis stratified by BMI (A), WC (B), WHtR (C), TyG-BMI (D), TyG-WC (E), and TyG-WHtR (F) groups. IE Infective Endocarditis; BMI Body Mass Index; WC Waist cicumference;WHtR Waist to height ratio; TyG Triglyceride-glucose

### Associations of Morphological and Metabolic Indices with Incident IE

Multivariable Cox proportional hazards models were utilized to evaluate the independent associations of the various indices with the risk of incident IE (Table 2). In the fully adjusted model (Model 3), all evaluated indices remained significantly associated with an elevated risk of IE. Notably, traditional morphological metrics reflecting central adiposity demonstrated comparatively greater predictive strength than both generalized obesity metrics and complex composite metabolic indices. Per 1-SD increment, WC yielded the highest risk estimate for incident IE (HR = 1.22, 95% CI: 1.14 – 1.31; P < 0.001), followed closely by WHtR (HR = 1.21, 95% CI: 1.14–1.29; P < 0.001).

Notably, indices incorporating the TyG index — an established surrogate for insulin resistance—were also significant independent predictors of IE, although their predictive strength was slightly more modest than that of isolated central adiposity metrics. Specifically, in the fully adjusted model, the HRs per 1-SD increment were 1.20 (95% CI: 1.12–1.28; P < 0.001) for TyG-WC and 1.19 (95% CI: 1.12–1.27; P < 0.001) for TyG-WHtR. Traditional BMI exhibited a hazard ratio of 1.19 (95% CI: 1.12 – 1.26). Furthermore, RCS analyses corroborated the robust, non-linear dose-response relationships between markers of central adiposity and IE risk (Figure 2).

**Fig. 2.**
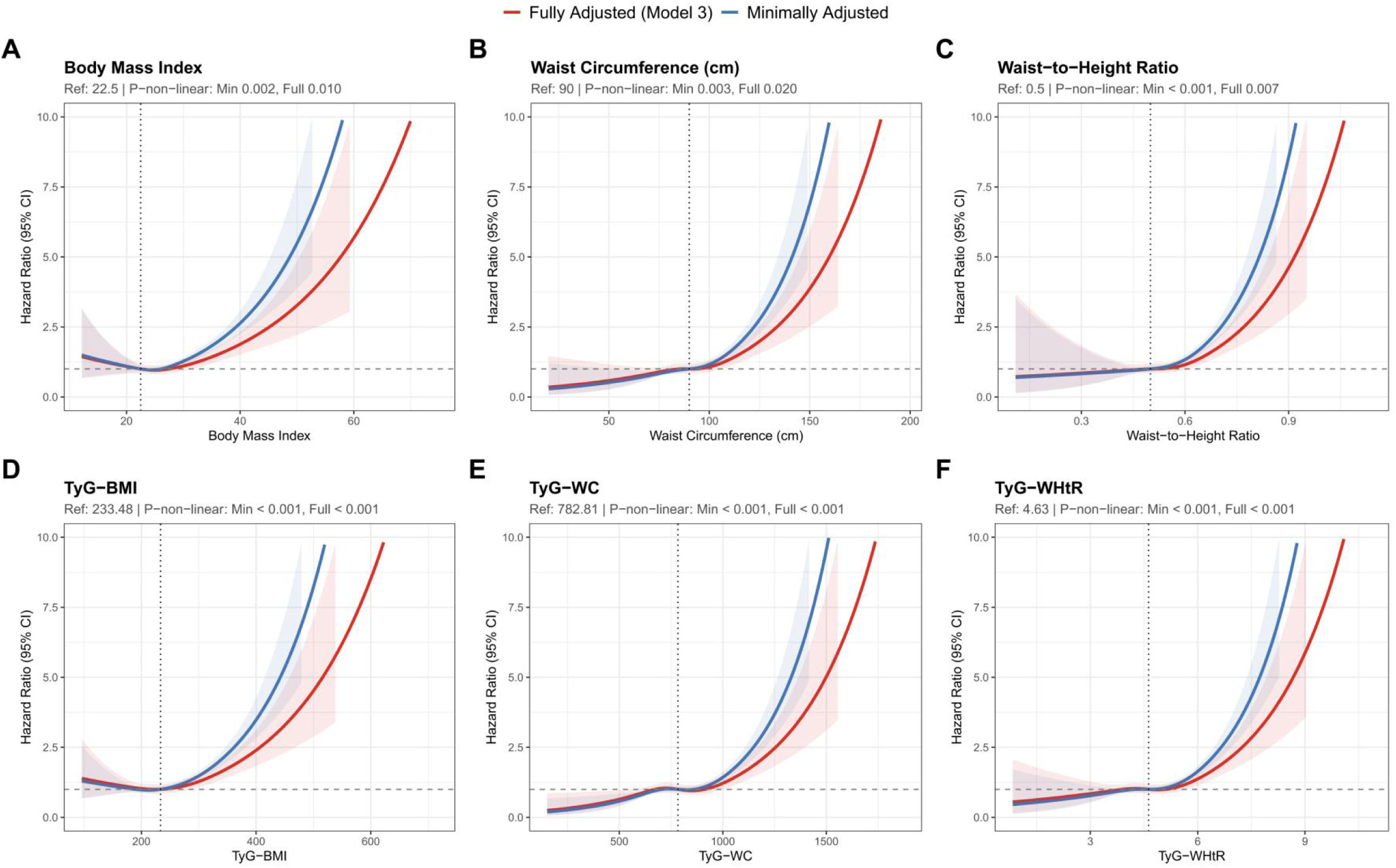
Restricted cubic spline curves for the dose-response relationships between BMI (A), WC (B), WHtR (C), TyG-BMI (D), TyG-WC (E), TyG-WHtR (F) and the risk of incident infective endocarditis. The minimally adjusted models (adjusted for age, sex, and race). The fully adjusted models (additionally adjusted for education, TDI, smoking, drinking, physical activity, and diabetes, hypertension, hypercholesterolemia, valvular heart disease, cardiovascular disease, and oral problems).

### Subgroup, Phenotypic, and Sensitivity Analyses

The stronger prognostic performance of central adiposity metrics remained consistent across major clinical subgroups (Figure 3). Significant effect modifications were observed for age and diabetes status; specifically, the magnitude of risk associated with increased WC was substantially greater among younger participants (< 65 years, HR = 1.25, 95% CI: 1.15–1.35; P < 0.001) and those with concomitant diabetes (HR = 1.41, 95% CI: 1.21 – 1.65; P < 0.001).Subgroup analysis results for BMI, TyG-BMI, TyG-WC, and TyG-WHtR are presented in the supplemental Figures S1–S4.

**Fig. 3.**
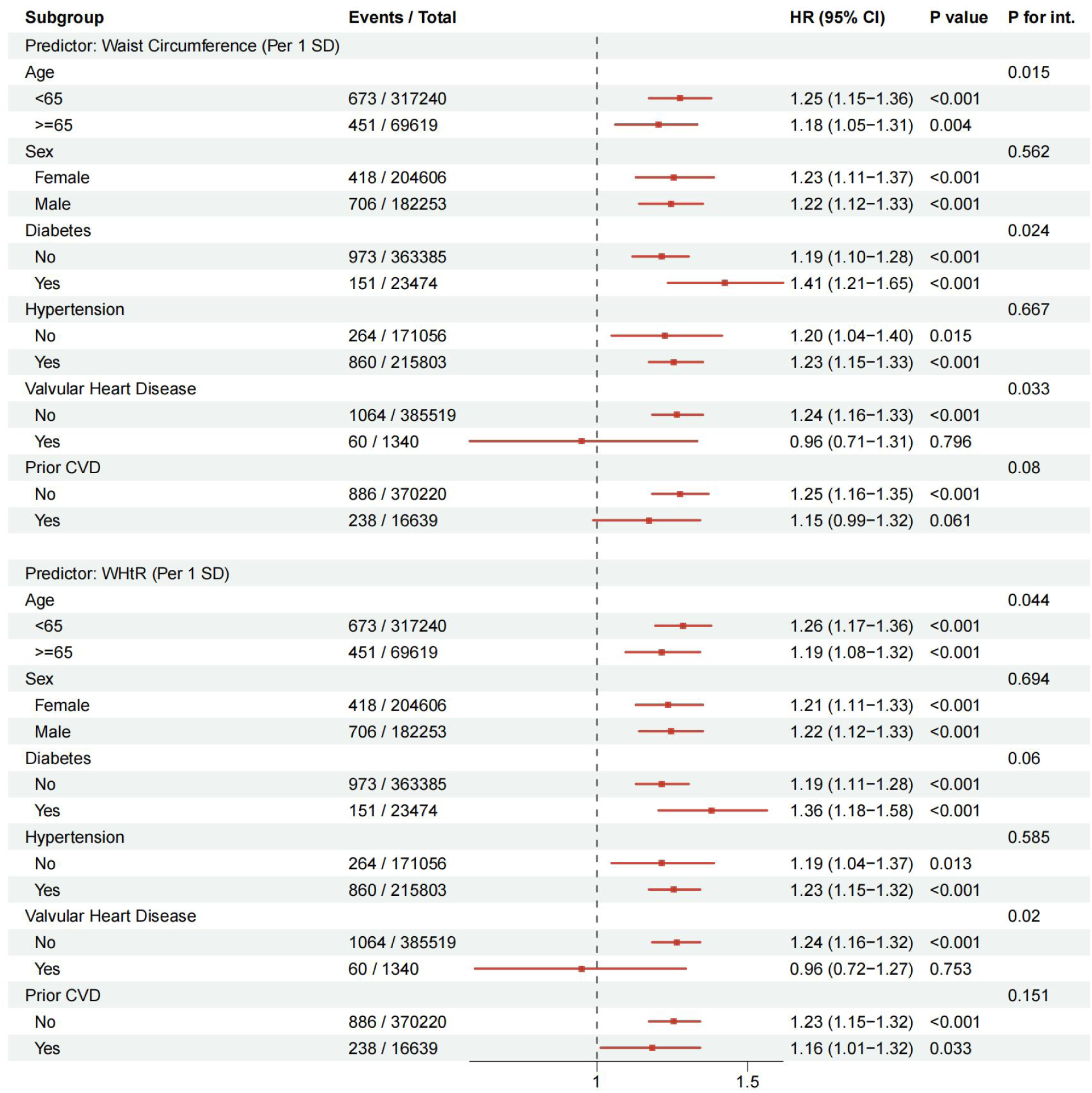
Forest plot of subgroup analyses of WC and WHtR for incident infective endocarditis. Adjusted for age, sex, race, education, Townsend deprivation index, smoking, drinking, physical activity, hypertension, diabetes, Hyperlipidemia, cardiovascular disease, valvular heart disease, oral problems. IE, infective endocarditis; WHtR, waist-to-height ratio.CVD, cardiovascular diseases;CI, confidence interval; HR, hazard ratio.

Upon stratifying the cohort into distinct metabolic-obesity phenotypes, the crude incidence of IE was highest in the MUO group (log-rank P < 0.001, Figure S5). Finally, rigorous sensitivity analyses — including Fine-Gray competing risk models, exclusion of individuals with valvular heart disease, and CCA — yielded virtually identical estimates (e.g., CCA HR for WC: 1.22, 95% CI: 1.14 – 1.31; P < 0.001). Moreover, the substantial E-values confirmed that our primary findings are highly robust against potential unmeasured confounding (Tables S5; S7;S8).

### Mediation Analysis for Systemic Inflammation

To further elucidate the pathophysiological pathways, we conducted a mediation analysis to evaluate the role of systemic inflammation (Table S6). The analysis revealed that the robust association between waist circumference and incident IE was partially, yet significantly, mediated by systemic inflammation. Specifically, the average causal mediation effect driven by CRP was highly significant (Estimate = -37.62, 95% CI: -69.82 to -9.98; P = 0.0076). Baseline CRP levels accounted for 20.76% (95% CI: 5.59% – 37.72%; P = 0.0076) of the total effect of waist circumference on IE risk. This empirical finding supports the hypothesis that visceral adiposity drives endocardial vulnerability, at least in part, through chronic pro-inflammatory pathways.

## Discussion

In this large-scale, prospective, population-based cohort study, we included 386,859 participants from the UK Biobank with a follow-up period spanning nearly 17 years. We comprehensively evaluated and compared the predictive value of traditional morphological indices versus complex metabolic markers for the risk of incident IE. Our primary finding was that traditional morphological indices — particularly WC and WHtR—were significantly and independently associated with IE risk. These indices outperformed BMI and insulin resistance-based metabolic markers, such as the TyG combined with WC (TyG-WC) or WHtR (TyG-WHtR). Notably, the risk associated with central obesity was more pronounced in individuals aged <65 years and those with concomitant diabetes mellitus. These associations remained statistically significant after adjusting for established IE risk factors.

To our knowledge, this study is the first to investigate the association between obesity-related metrics and IE risk. Previous literature has established that obesity directly contributes to cardiovascular risk factors, and central obesity—determined by WC and WHtR — is a cardiovascular risk factor independent of BMI^6,32,33^. While traditional epidemiological understanding suggests that IE is primarily secondary to structural heart disease, our 16-year follow-up confirms that morphological alterations alone can independently drive IE risk. Our findings align with a study of Swedish adolescents by Sourander et al., which identified a strong, dose-dependent association between high BMI and IE^34^. Furthermore, a multicenter analysis of the German CAMPAIGN registry involving 4,801 IE patients undergoing cardiac surgery revealed that obese patients (BMI ≥ 30 kg/m ²) carried a higher burden of comorbidities, including hypertension (63.0%), diabetes (45.7%), and coronary artery disease (29.5%). That study also reported the highest prevalence of staphylococcal endocarditis (33.4%) in the obese group, with 30-day (14.1%) and 1-year (19.6%) mortality rates significantly exceeding those of other weight groups^35^. These observations suggest that obesity not only increases IE incidence but may also worsen prognosis by altering the pathogen spectrum and increasing comorbidity burden.

Although BMI is widely used as a core metric for obesity, it fails to differentiate between adipose distribution and muscle mass. Research by Frías-García et al. indicated that IE patients with low body weight (BMI ≤ 18.5 kg/m ²) had a significantly higher 1-year mortality (HR 1.94, 95% CI 1.14–3.31), suggesting that underweight status is also a predictor of adverse outcomes^36^. In contrast, central obesity indices capture visceral fat accumulation — a critical metabolic phenotype. Visceral fat may contribute to IE pathogenesis through mechanisms such as chronic low-grade inflammation and endothelial dysfunction^37,38^.This physiological framework is strongly corroborated by our mediation analysis, which demonstrated that over 20% of the pro-infective effect of waist circumference is directly attributable to elevated systemic inflammation (CRP).

Stratified analysis by metabolic health status showed that, compared with MHNW individuals, both MHO (HR 1.23, P = 0.03) and MUO (HR 1.27, P = 0.008) statuses were independently associated with increased IE risk. Conversely, the MUNW status did not reach statistical significance (HR 1.06, P = 0.637). These results indicate that obesity itself — regardless of metabolic status — is the primary driver of IE risk, whereas isolated metabolic abnormalities in normal-weight individuals appear insufficient to significantly increase risk. This supports our overarching conclusion that morphological changes alone are independent drivers of IE.

Interestingly, we observed that the surge in IE risk associated with central obesity and metabolic abnormalities was more acute in individuals aged <65 years. For instance, each standard deviation increase in WC was associated with a 25% risk increase in those <65 years, compared to only 17% in those ≥ 65 years. This phenomenon may be attributed to age-related competing risks. The study noted a median age of 65 for IE patients, and advanced age itself is a strong independent risk factor that may partially mask the relative contribution of obesity^35^. Furthermore, obesity in younger populations often represents a “purer” phenotype with fewer age-related comorbidities, making its pathogenic effects more apparent. From a preventive perspective, this suggests that weight intervention in young and middle-aged populations may yield greater absolute benefits.

A nationwide cohort study by Kim et al. involving 1,762,108 Korean patients with diabetes revealed a J-shaped association between weight change and IE risk. Compared to the stable weight group, the hazard ratio for IE was 2.41 (95% CI 1.87– 3.12) in the severe weight loss group (≤-10%) and 1.59 (95% CI 1.11–2.28) in the severe weight gain group ( ≥ 10%)^39^. This suggests that patients with diabetes are particularly sensitive to weight fluctuations. Both obesity and emaciation may increase IE risk through distinct mechanisms. In patients with diabetes, impaired immune function, compromised skin and mucosal barriers, and a hyperglycemic environment conducive to pathogen growth may act synergistically with obesity-related chronic inflammation to increase susceptibility to valvular infection.

Among patients with pre-existing valvular heart disease, the HRs for all obesity-related indices fell to the 0.89 – 0.96 range and lost statistical significance. This likely reflects a “ceiling effect” in etiology; the pathogenic effect of structural heart disease is so dominant that the relative contribution of obesity, a comparatively modest risk factor, becomes undetectable in additive analyses. While IE is traditionally viewed as secondary to structural heart disease, our 16-year follow-up demonstrates that in populations without structural heart disease, morphological changes can independently drive IE risk. The null results in the valvular disease subgroup further validate this: when a potent risk factor like structural pathology is present, the relative impact of metabolic risk factors is diluted.

This study possesses several strengths, including its prospective design, large sample size, and a follow-up period of nearly 17 years (exceeding 6.3 million person-years), providing robust statistical power. Additionally, the use of restricted cubic splines elucidated non-linear dose-response relationships, while Fine-Gray models was employed to control for competing risks and unmeasured confounding. However, several limitations warrant consideration. First, the observational design precludes definitive causal inference. Second, UK Biobank participants are predominantly of European descent and subject to “healthy volunteer” selection bias, which may limit the generalizability of these findings to other ethnicities or broader populations. Third, our analysis relied on a single baseline measurement, preventing an assessment of dynamic changes in obesity and metabolic status during follow-up.

## Conclusion

In conclusion, central adiposity, specifically as assessed by waist circumference and waist-to-height ratio, is a robust and independent risk factor for incident infective endocarditis. These simple anthropometric measures demonstrate superior prognostic value compared to traditional BMI and complex metabolic composites, such as TyG-related indices.

## Abbreviations

BMI: body mass index
CCA: complete-case analyses
CI: confidence interval
CRP: C-reactive protein
DBP: diastolic blood pressure
FBG: fasting blood glucose
HbA1c: hemoglobin A1c
HDL-C: high-density lipoprotein-cholesterol
HR: hazard ratios
ICD-10: International Classification of Disease versions 10
IE: Infective Endocarditis
IQR: interquartile range
IR: insulin resistance
LDL-C: low-density lipoprotein cholesterol
MET: metabolic equivalent of task
MHNW: metabolically healthy normal weight
MHO: metabolically healthy obesity
MUNW: metabolically unhealthy normal weight
MUO: metabolically unhealthy obesity.
ROC: receiver operating characteristic
RCS: restricted cubic spline
SBP: systolic blood pressure
SD: standard deviation
TC: total cholesterol
TDI: Townsend deprivation index
TG: triglycerides
TyG: triglyceride-glucose
WC: waist circumference
WHtR: waist-to-height ratio

## Ethics approval and consent to participate

Ethics approval and consent to participate UK Biobank has been approved by the National Research Ethics Committee of the National Health Service (NHS). All participants signed informed consent.

## Consent for publication

Not applicable.

## Availability of data and materials

Researchers interested in accessing the data used in this study can apply for access to the UK Biobank by visiting their website (https://www.ukbiobank.ac.uk/enable-your-research/apply-for-access) and submitting an application that includes a research protocol summary and requested data fields. Upon approval by the UK Biobank management team and payment of applicable fees, researchers will be granted access to the dataset.

## Author contributions

Changwei Ren,Song Wang designed the research; Song Wang and Jinwei Zhang performed the statistical analysis; Song Wang, Peng Sun, Zhipeng Wei, Ke Zhang, Chenzhen Xu, Chuanjie Yue, Yuzhe Zhang Lingyao Li, Liaoming He and Jianbo Yu contributed to data collection and interpretation; Song Wang wrote the paper; Changwei Ren, Hao Cui, Yongqiang Lai revised the manuscript. All authors read and approved the final manuscript.

## Acknowledgements

This study has been conducted using the UK Biobank Resources under application number 545415.

---

**Table S1.**
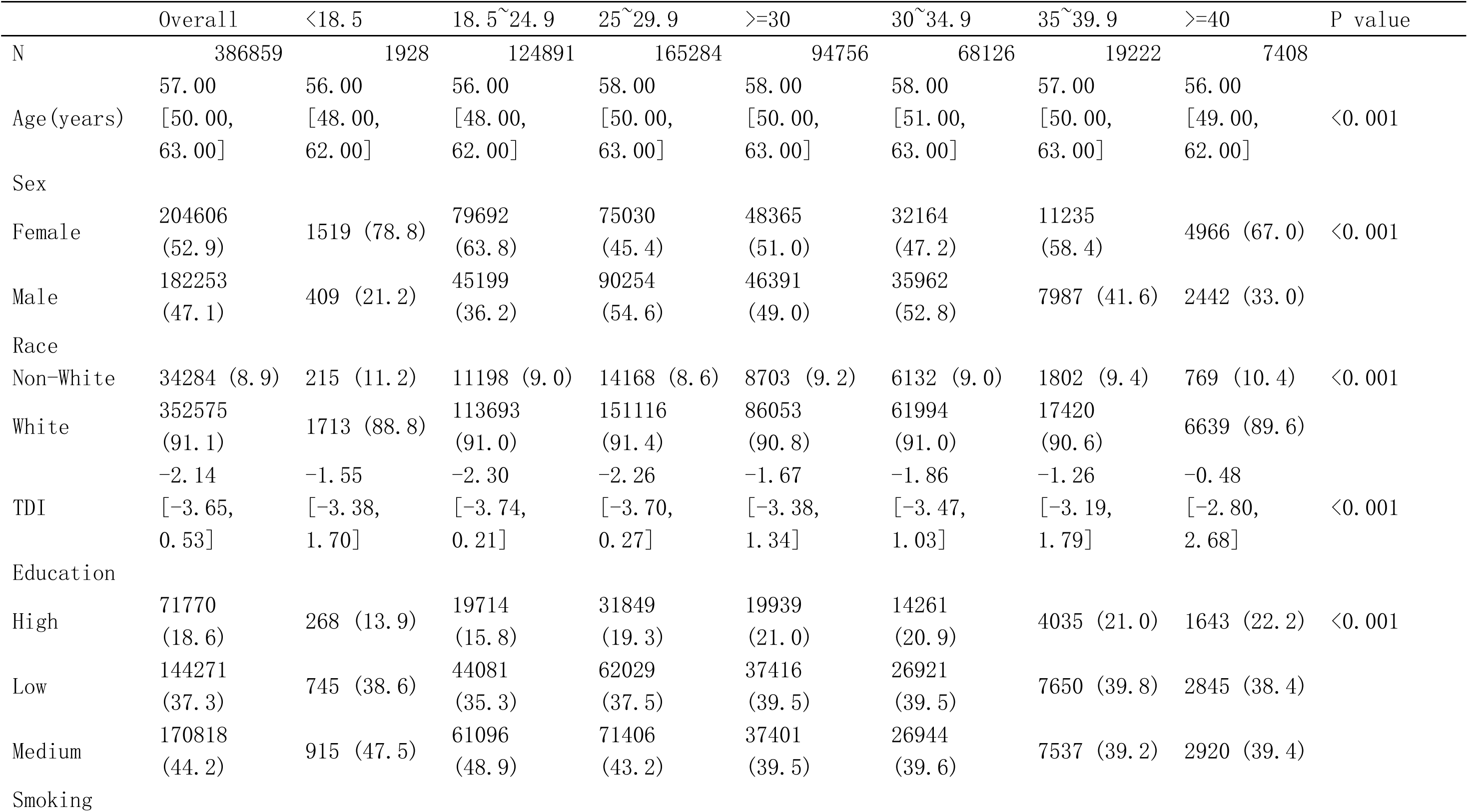
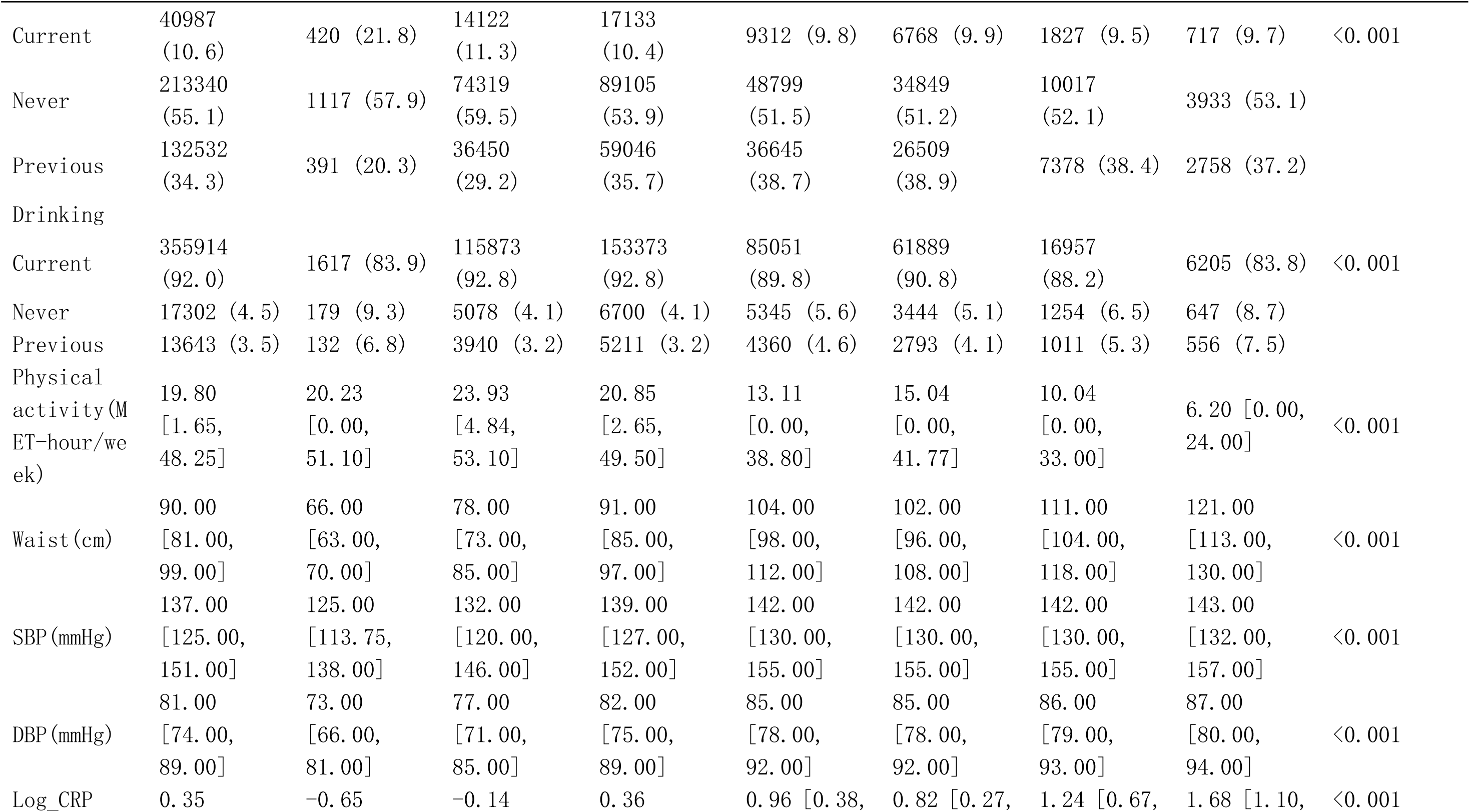
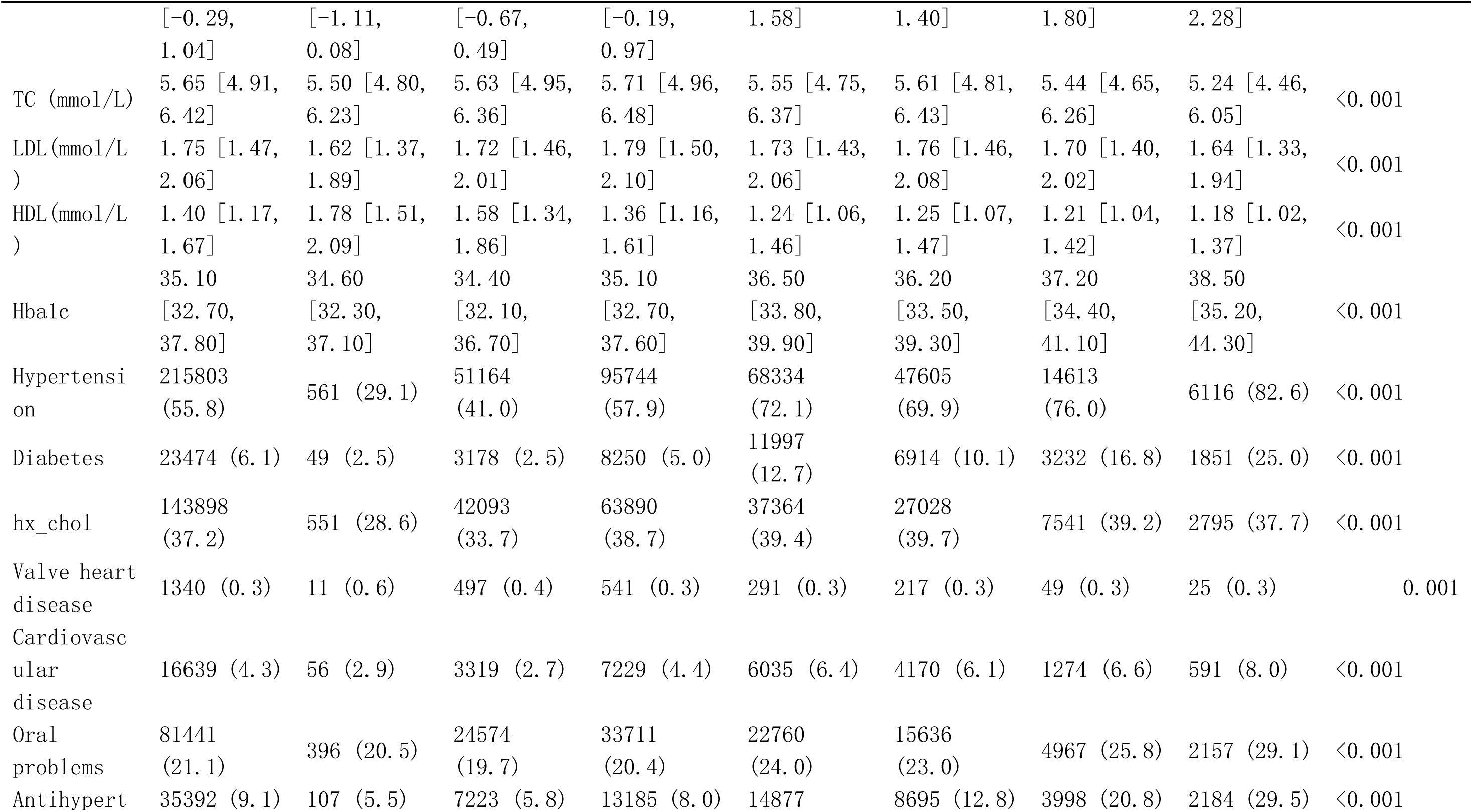
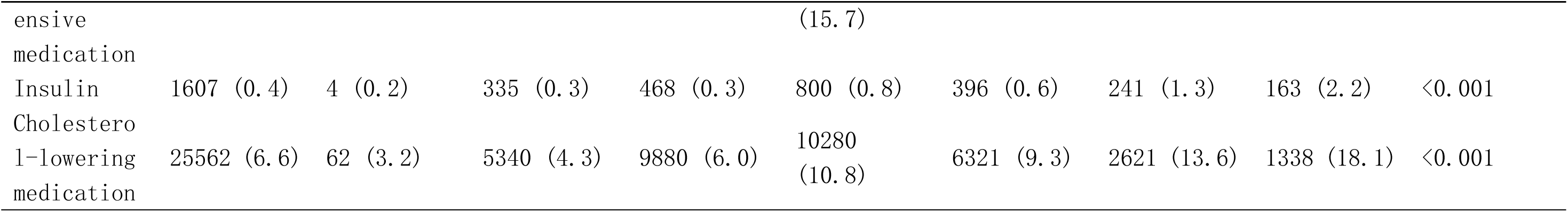
Baseline characteristics of the study individuals according to BMI quartiles.

**Table S2.**
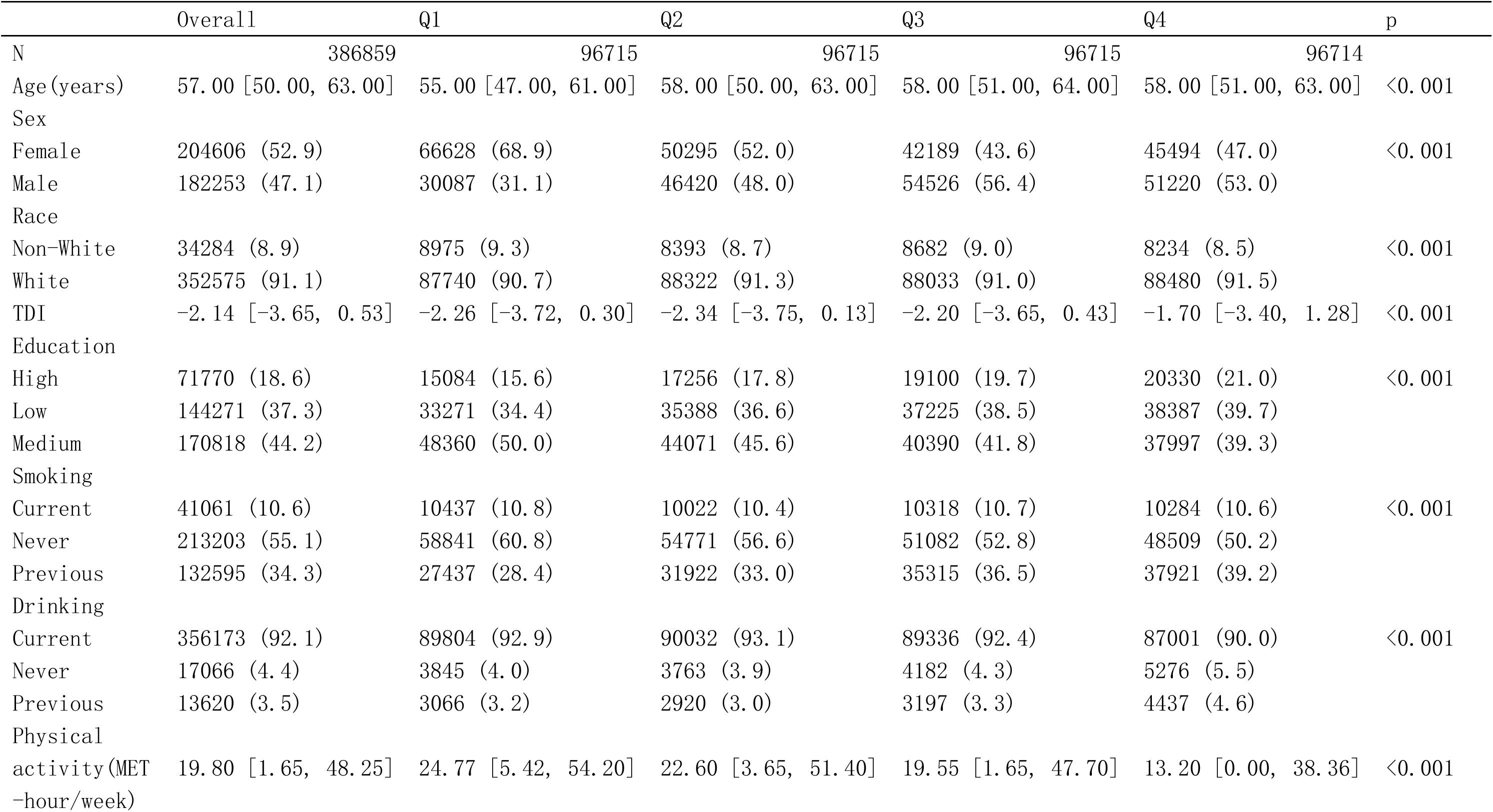
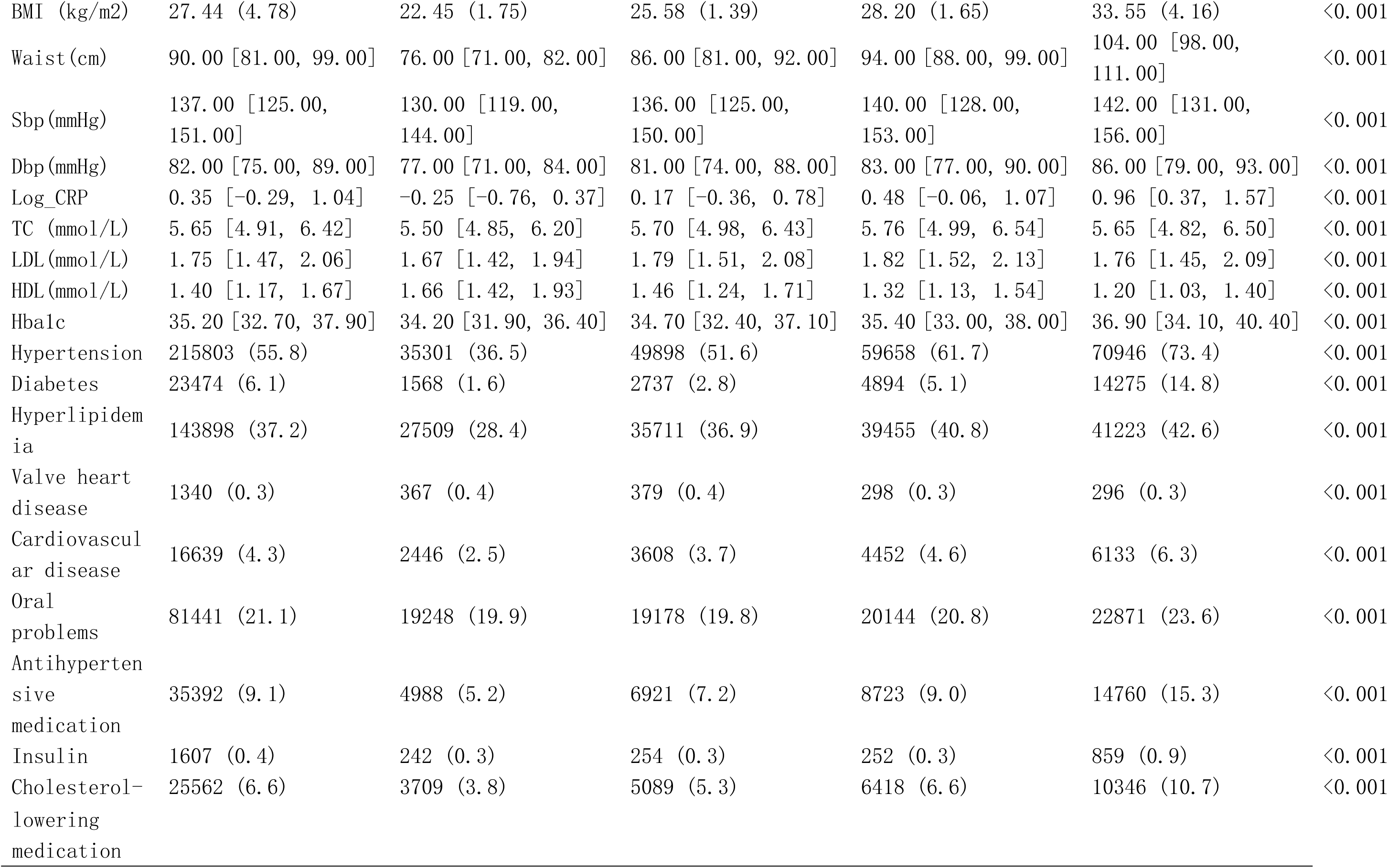
Baseline characteristics of the study individuals according to TyG-BMI quartiles.

**Table S3.**
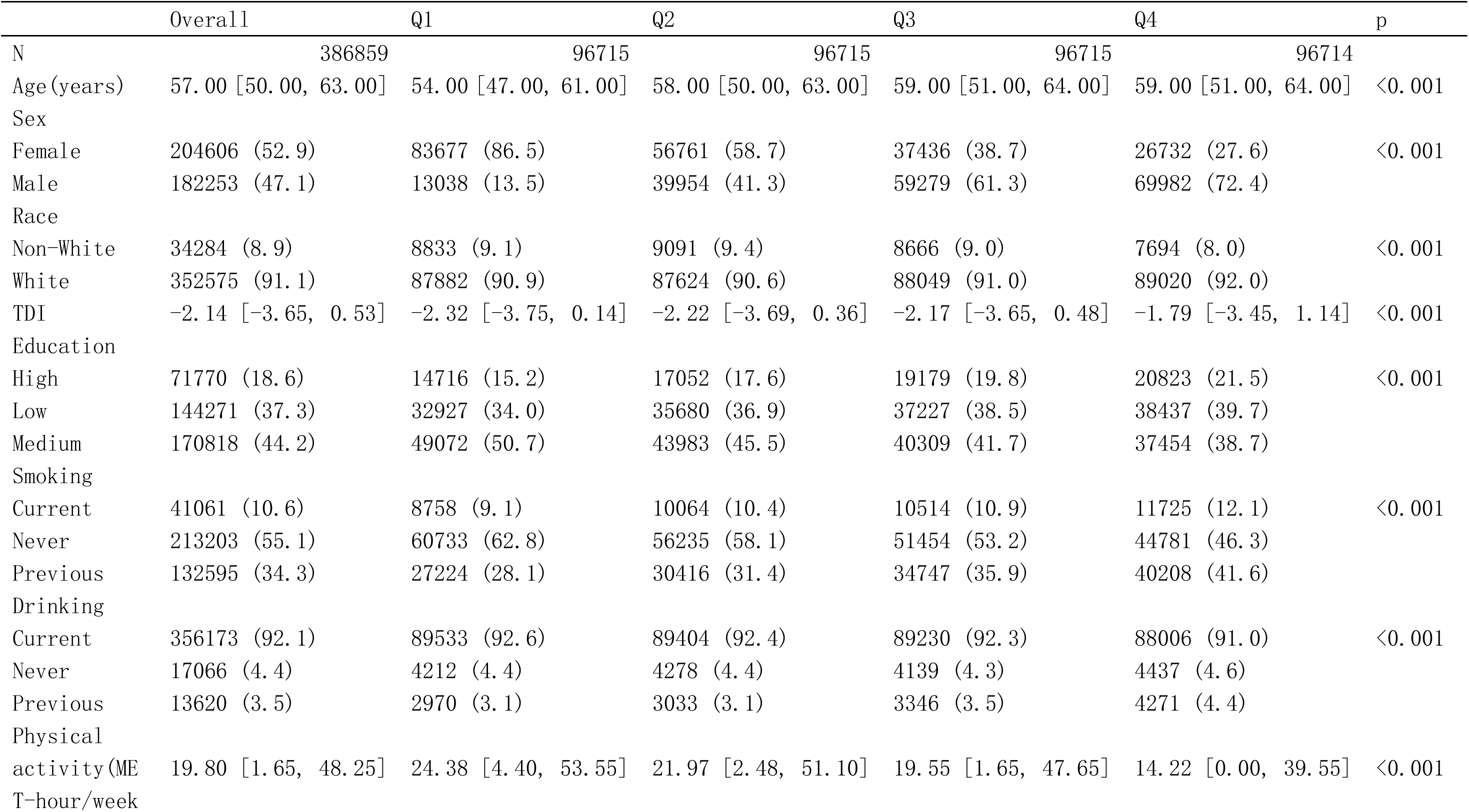
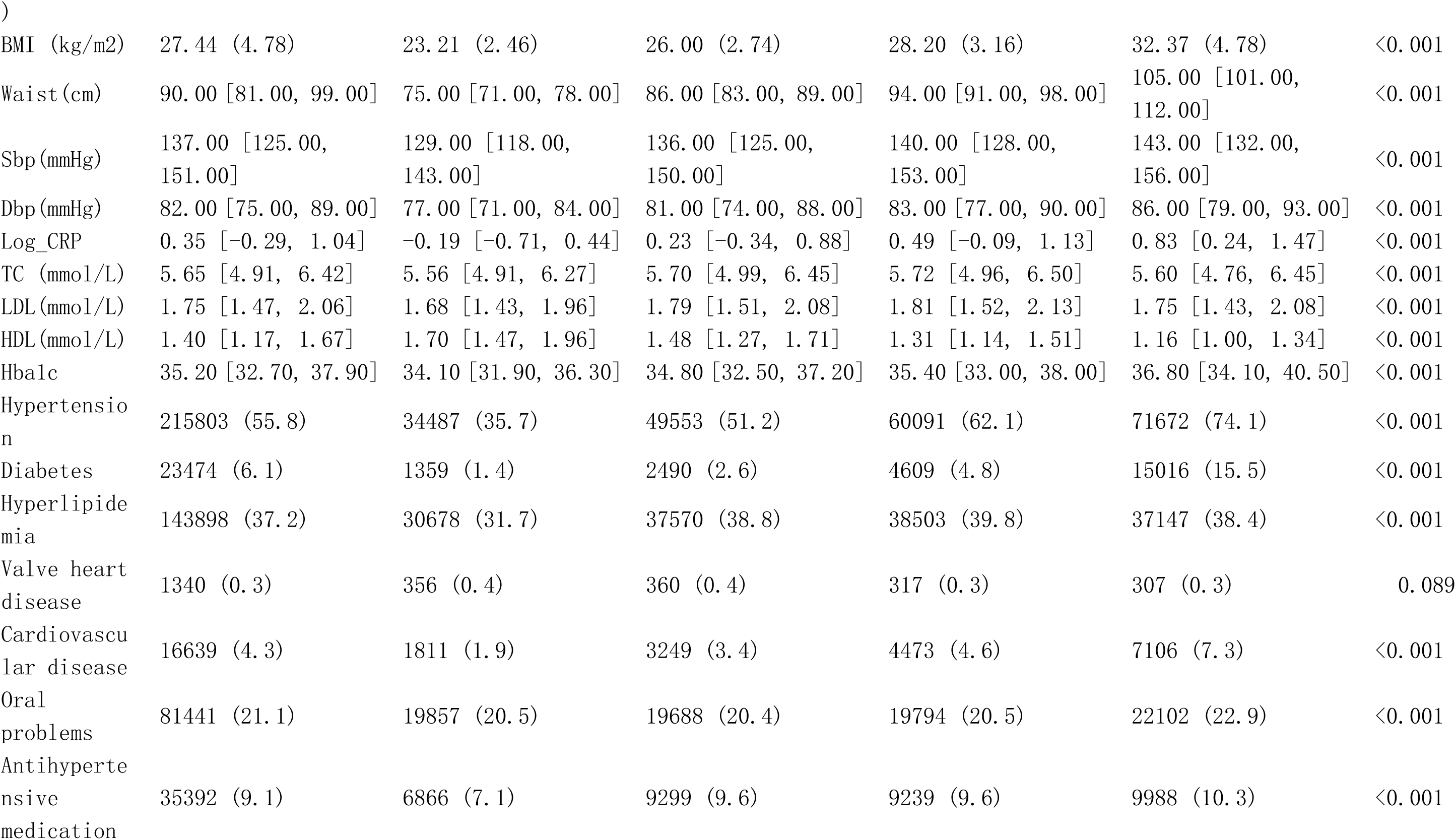
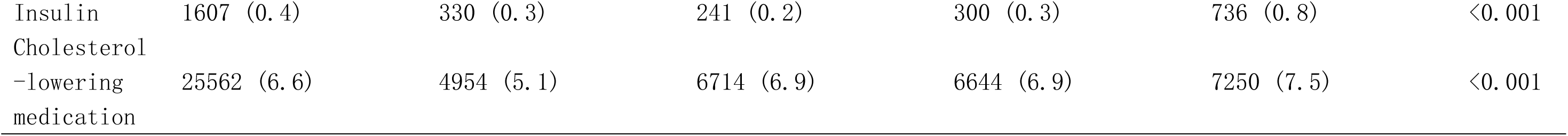
Baseline characteristics of the study individuals according to TyG-WC quartiles.

**Table S4.**
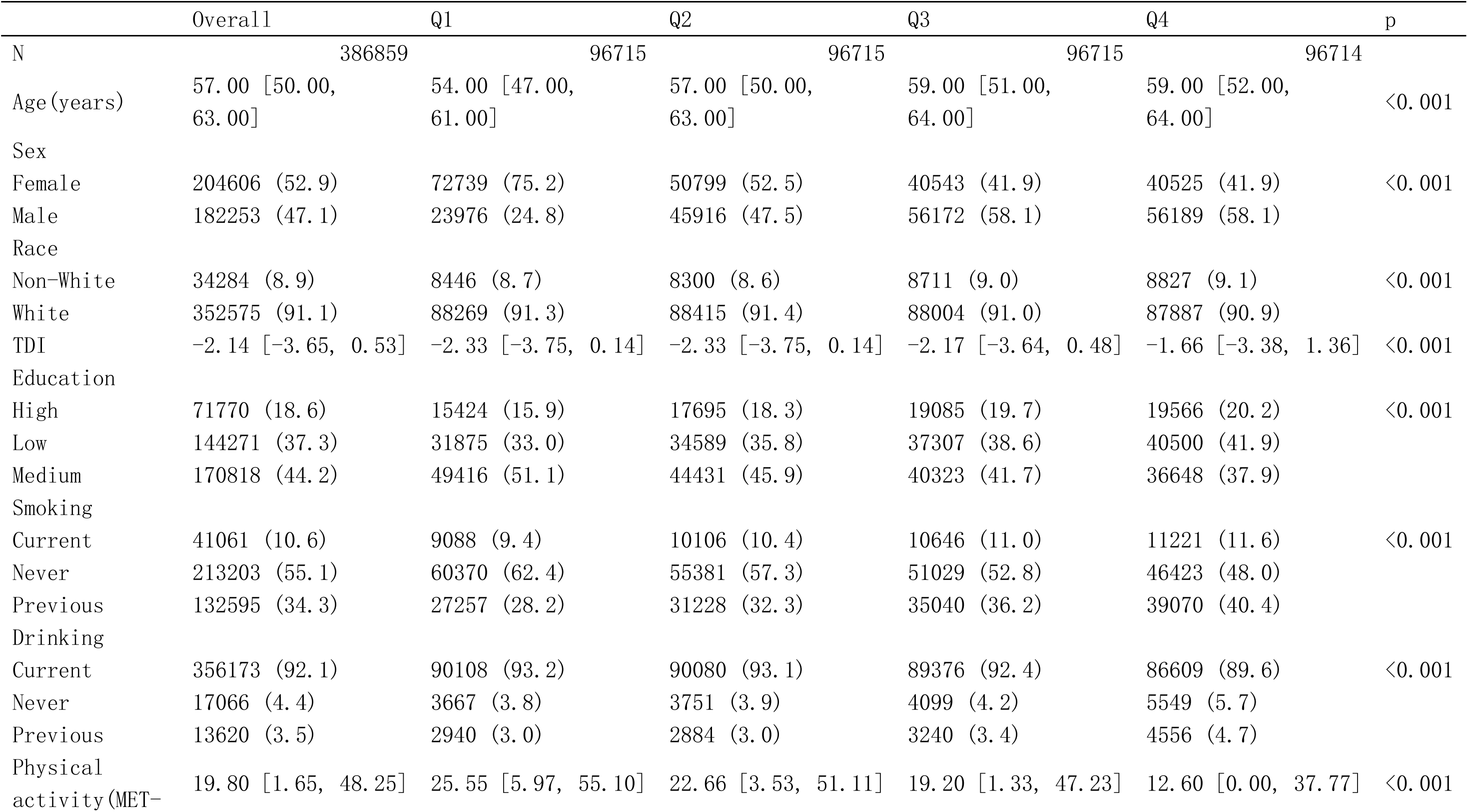
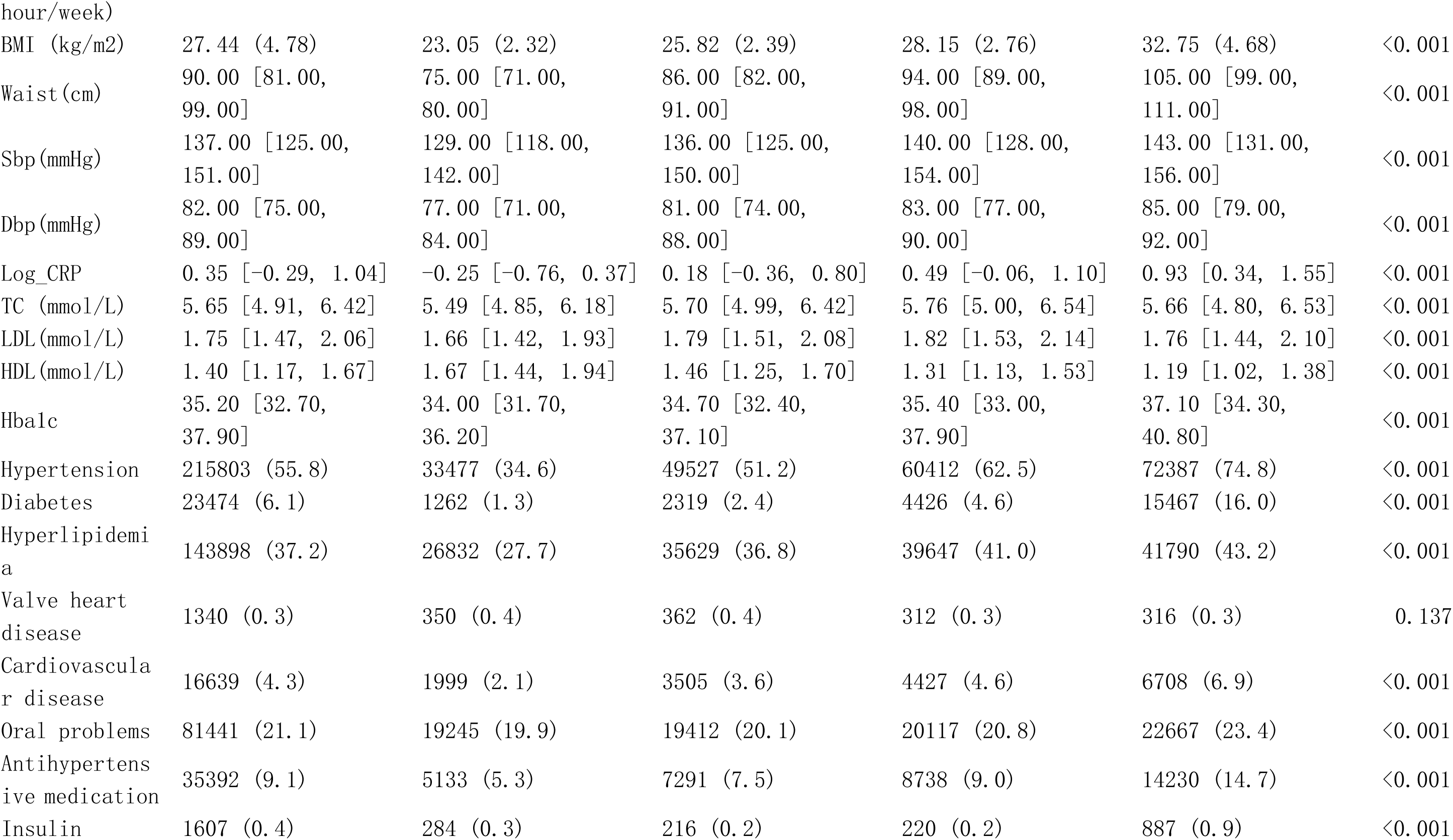
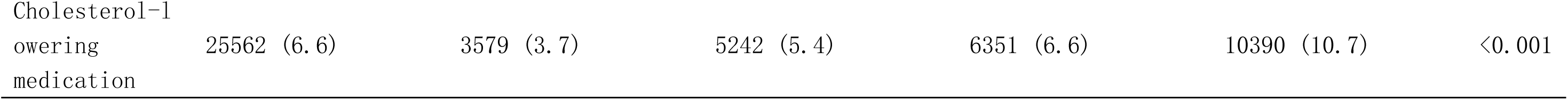
Baseline characteristics of the study individuals according to TyG-WHtR quartiles.

**Table S5.** Sensitivity analyses for the associations of WC, WHtR, and TyG-related indices with incident infective endocarditis.

| Variable | row_names | M1_HR | M1_P | M2_HR | M2_P | M3_HR | M3_P |
| --- | --- | --- | --- | --- | --- | --- | --- |
| BMI | <18.5 | 1.02 (0.38–2.73) | 0.972 | 1.13 (0.42–3.03) | 0.814 | 1.18 (0.44–3.17) | 0.741 |
|  | 18.5~24.9 | Reference |  | Reference |  | Reference |  |
|  | 25~29.9 | 1.27 (1.09–1.47) | 0.002 | 1.01 (0.86–1.17) | 0.937 | 0.96 (0.82–1.12) | 0.566 |
|  | 30~34.9 | 1.88 (1.59–2.23) | <0.001 | 1.48 (1.24–1.75) | <0.001 | 1.29 (1.08–1.54) | 0.005 |
|  | 35~39.9 | 2.36 (1.87–2.99) | <0.001 | 2.04 (1.61–2.59) | <0.001 | 1.65 (1.29–2.11) | <0.001 |
|  | >=40 | 2.79 (2.01–3.86) | <0.001 | 2.77 (2.00–3.85) | <0.001 | 2.06 (1.47–2.89) | <0.001 |
|  | P for trend (Main) |  | <0.001 |  | <0.001 |  | <0.001 |
|  | P for trend (Sub) |  | 0.976 |  | 0.77 |  | 0.741 |
|  | Per 1 kg/m2 | 1.06 (1.05–1.07) | <0.001 | 1.05 (1.04–1.07) | <0.001 | 1.04 (1.03–1.05) | <0.001 |
|  | Per SD | 1.31 (1.24–1.37) | <0.001 | 1.28 (1.21–1.35) | <0.001 | 1.19 (1.13–1.27) | <0.001 |
| Waist | Q1 | Reference |  | Reference |  | Reference |  |
|  | Q2 | 1.63 (1.33–2.01) | <0.001 | 1.24 (1.00–1.53) | 0.051 | 1.21 (0.97–1.50) | 0.084 |
|  | Q3 | 2.15 (1.76–2.62) | <0.001 | 1.38 (1.11–1.70) | 0.003 | 1.28 (1.03–1.59) | 0.024 |
|  | Q4 | 3.17 (2.63–3.83) | <0.001 | 1.83 (1.48–2.26) | <0.001 | 1.54 (1.24–1.91) | <0.001 |
|  | P for trend |  | <0.001 |  | <0.001 |  | <0.001 |
|  | Per 1 cm | 1.03 (1.03–1.04) | <0.001 | 1.02 (1.02–1.03) | <0.001 | 1.02 (1.01–1.02) | <0.001 |
|  | Per SD | 1.53 (1.45–1.61) | <0.001 | 1.33 (1.25–1.42) | <0.001 | 1.23 (1.15–1.32) | <0.001 |
| WHtR | Q1 | Reference |  | Reference |  | Reference |  |
|  | Q2 | 1.55 (1.27–1.89) | <0.001 | 1.13 (0.92–1.38) | 0.238 | 1.10 (0.90–1.36) | 0.339 |
|  | Q3 | 1.66 (1.36–2.02) | <0.001 | 1.01 (0.83–1.24) | 0.901 | 0.95 (0.78–1.17) | 0.655 |
|  | Q4 | 3.04 (2.54–3.64) | <0.001 | 1.73 (1.43–2.09) | <0.001 | 1.46 (1.20–1.78) | <0.001 |
|  | P for trend |  | <0.001 |  | <0.001 |  | <0.001 |
|  | Per 1 unit | 212.63 (106.35–425.10) | <0.001 | 38.79 (17.60–85.52) | <0.001 | 14.64 (6.38–33.61) | <0.001 |
|  | Per SD | 1.49 (1.42–1.57) | <0.001 | 1.31 (1.24–1.39) | <0.001 | 1.22 (1.15–1.30) | <0.001 |
| TyG-BMI | Q1 | Reference |  | Reference |  | Reference |  |
|  | Q2 | 1.29 (1.07–1.56) | 0.008 | 0.98 (0.81–1.19) | 0.846 | 0.95 (0.78–1.15) | 0.582 |
|  | Q3 | 1.41 (1.17–1.70) | <0.001 | 0.97 (0.81–1.17) | 0.779 | 0.91 (0.76–1.10) | 0.343 |
|  | Q4 | 2.12 (1.78–2.51) | <0.001 | 1.49 (1.25–1.78) | <0.001 | 1.25 (1.04–1.50) | 0.017 |
|  | P for trend |  | <0.001 |  | <0.001 |  | 0.007 |
|  | Per 1 unit | 1.01 (1.00–1.01) | <0.001 | 1.00 (1.00–1.01) | <0.001 | 1.00 (1.00–1.00) | <0.001 |
|  | Per SD | 1.34 (1.28–1.41) | <0.001 | 1.27 (1.21–1.35) | <0.001 | 1.19 (1.12–1.26) | <0.001 |
| TyG-WC | Q1 | Reference |  | Reference |  | Reference |  |
|  | Q2 | 1.92 (1.57–2.35) | <0.001 | 1.36 (1.10–1.67) | 0.004 | 1.31 (1.07–1.62) | 0.01 |
|  | Q3 | 1.91 (1.56–2.34) | <0.001 | 1.13 (0.91–1.40) | 0.257 | 1.07 (0.86–1.33) | 0.567 |
|  | Q4 | 3.18 (2.63–3.84) | <0.001 | 1.72 (1.39–2.11) | <0.001 | 1.44 (1.16–1.79) | <0.001 |
|  | P for trend |  | <0.001 |  | <0.001 |  | 0.007 |
|  | Per 1 unit | 1.00 (1.00–1.00) | <0.001 | 1.00 (1.00–1.00) | <0.001 | 1.00 (1.00–1.00) | <0.001 |
|  | Per SD | 1.51 (1.43–1.59) | <0.001 | 1.30 (1.22–1.38) | <0.001 | 1.20 (1.12–1.29) | <0.001 |
| TyG-WHtR | Q1 | Reference |  | Reference |  | Reference |  |
|  | Q2 | 1.64 (1.35–2.00) | <0.001 | 1.14 (0.93–1.39) | 0.201 | 1.10 (0.90–1.35) | 0.34 |
|  | Q3 | 1.76 (1.45–2.14) | <0.001 | 1.04 (0.85–1.27) | 0.697 | 0.98 (0.80–1.21) | 0.868 |
|  | Q4 | 2.84 (2.37–3.41) | <0.001 | 1.56 (1.29–1.88) | <0.001 | 1.31 (1.07–1.60) | 0.008 |
|  | P for trend |  | <0.001 |  | <0.001 |  | 0.009 |
|  | Per 1 unit | 1.61 (1.51–1.72) | <0.001 | 1.36 (1.26–1.46) | <0.001 | 1.25 (1.15–1.35) | <0.001 |
|  | Per SD | 1.49 (1.41–1.57) | <0.001 | 1.29 (1.21–1.37) | <0.001 | 1.20 (1.13–1.28) | <0.001 |

**Table S6.** Mediation analyses of the associations between waist circumference and incident infective endocarditis.

| Effect Path | Estimate | 95% CI | P Value | Proportion Mediated (%) |
| --- | --- | --- | --- | --- |
| Total Effect | -193.60135 | -287.48727 - -118.78792 | P<0.001 | - |
| Average Direct Effect (ADE) | -155.13218 | -241.34449 - -84.37046 | P<0.001 | - |
| Average Causal Mediation Effect (ACME) | -38.46918 | -70.70637 - -11.28830 | P=0.0072 | - |
| Proportion Mediated | 20.70% | 6.42% - 37.67% | P=0.0076 | 20.70% |

**Table S7.** Associations of BMI, WC, WHtR, and TyG-related indices with incident infective endocarditis considering death as a competing risk.

| Predictor | Model | SHR_95CI | P_value |
| --- | --- | --- | --- |
| bmi | Fine-Gray<br>(Competing Risk) | 1.18<br>(1.12–1.26) | <0.001 |
| waist | Fine-Gray<br>(Competing Risk) | 1.21<br>(1.13–1.30) | <0.001 |
| whtr | Fine-Gray<br>(Competing Risk) | 1.21<br>(1.13–1.29) | <0.001 |
| TyG_BMI | Fine-Gray<br>(Competing Risk) | 1.18<br>(1.11–1.26) | <0.001 |
| TyG_WC | Fine-Gray<br>(Competing Risk) | 1.19<br>(1.11–1.28) | <0.001 |
| TyG_WHtR | Fine-Gray<br>(Competing Risk) | 1.19<br>(1.11–1.27) | <0.001 |

**Table S8.**
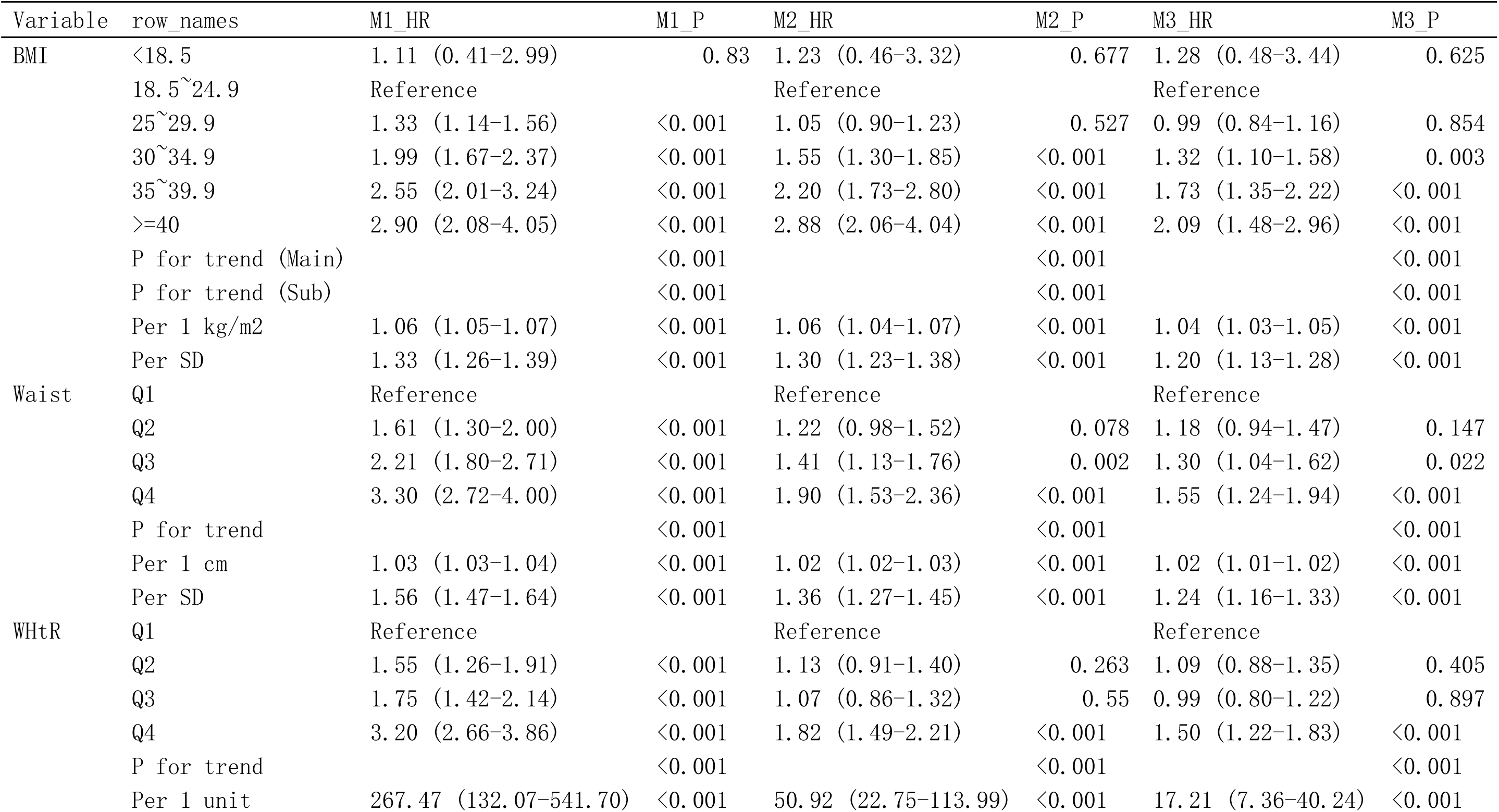
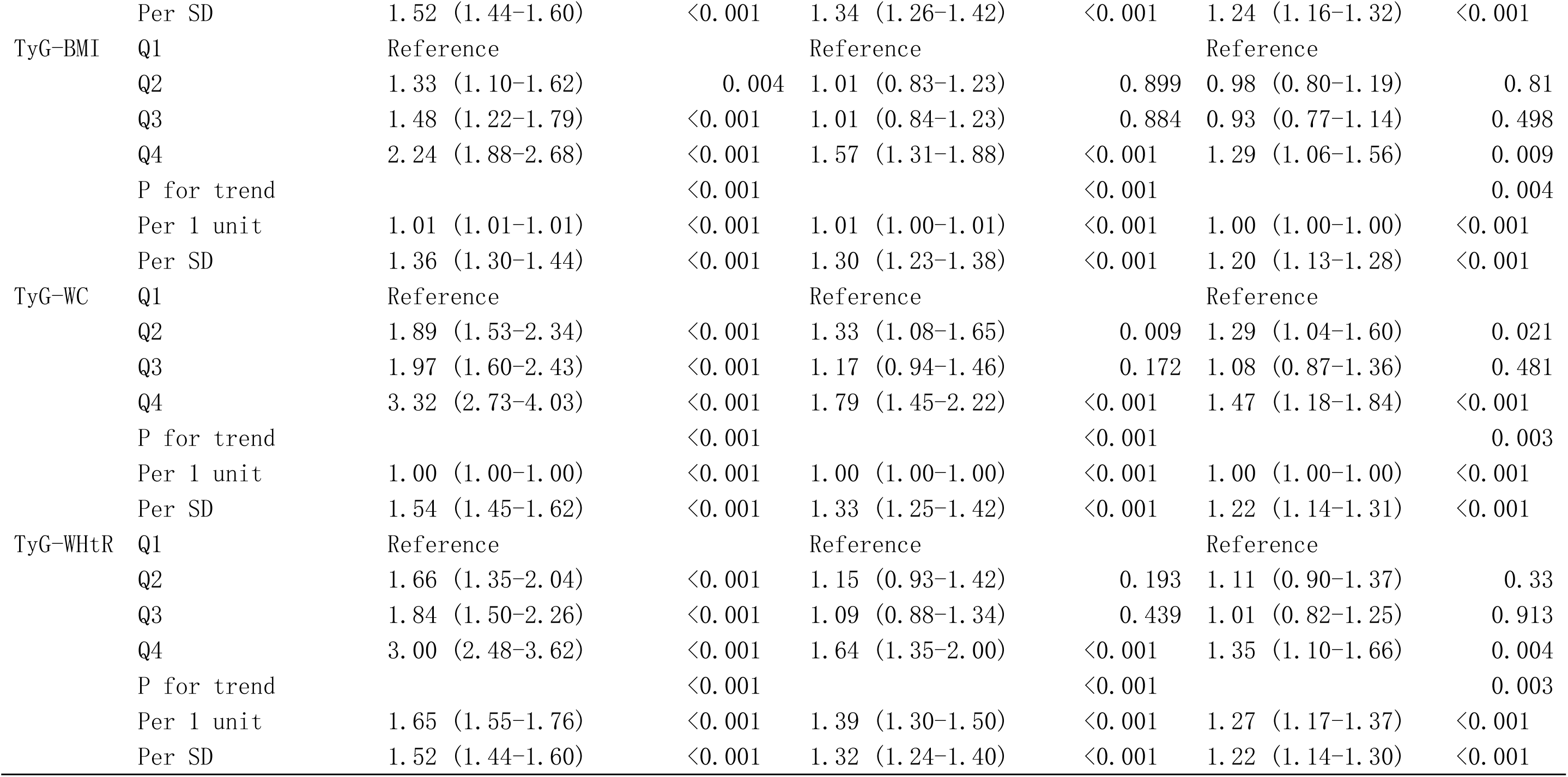
Associations of WC, WHtR, and TyG-related indices with incident infective endocarditis after excluding individuals with valvular heart disease.

**Table S9.** Baseline characteristics of the study individuals according to waist-to-height ratio quartiles.

| Variables | Overall | Q1 | Q2 | Q3 | Q4 | P value |
| --- | --- | --- | --- | --- | --- | --- |
| N | 386859 | 96715 | 96715 | 96715 | 96714 |  |
| Age(years) | 57.00 [50.00, 63.00] | 54.00 [47.00, 61.00] | 57.00 [49.00, 63.00] | 59.00 [51.00, 64.00] | 59.00 [52.00, 64.00] | <0.001 |
| **Sex** |  |  |  |  |  |  |
| Female | 204606 (52.9) | 72826 (75.3) | 48879 (50.5) | 39318 (40.7) | 43583 (45.1) | <0.001 |
| Male | 182253 (47.1) | 23889 (24.7) | 47836 (49.5) | 57397 (59.3) | 53131 (54.9) |  |
| **Race** |  |  |  |  |  |  |
| Non-White | 34284 (8.9) | 7858 (8.1) | 8272 (8.6) | 8703 (9.0) | 9451 (9.8) | <0.001 |
| White | 352575 (91.1) | 88857 (91.9) | 88443 (91.4) | 88012 (91.0) | 87263 (90.2) |  |
| TDI | -2.14 [-3.65, 0.53] | -2.35 [-3.77, 0.09] | -2.35 [-3.76, 0.11] | -2.17 [-3.65, 0.44] | -1.57 [-3.34, 1.47] | <0.001 |
| **Education** |  |  |  |  |  |  |
| High | 71770 (18.6) | 15126 (15.6) | 17915 (18.5) | 19298 (20.0) | 19431 (20.1) | <0.001 |
| Low | 144271 (37.3) | 32135 (33.2) | 34247 (35.4) | 36885 (38.1) | 41004 (42.4) |  |
| Medium | 170818 (44.2) | 49454 (51.1) | 44553 (46.1) | 40532 (41.9) | 36279 (37.5) |  |
| **Smoking** |  |  |  |  |  |  |
| Current | 41061 (10.6) | 9707 (10.0) | 10335 (10.7) | 10501 (10.9) | 10518 (10.9) | <0.001 |
| Never | 213203 (55.1) | 60114 (62.2) | 55315 (57.2) | 50592 (52.3) | 47182 (48.8) |  |
| Previous | 132595 (34.3) | 26894 (27.8) | 31065 (32.1) | 35622 (36.8) | 39014 (40.3) |  |
| **Drinking** |  |  |  |  |  |  |
| Current | 356173 (92.1) | 90001 (93.1) | 90244 (93.3) | 89555 (92.6) | 86373 (89.3) | <0.001 |
| Never | 17066 (4.4) | 3739 (3.9) | 3626 (3.7) | 4021 (4.2) | 5680 (5.9) |  |
| Previous | 13620 (3.5) | 2975 (3.1) | 2845 (2.9) | 3139 (3.2) | 4661 (4.8) |  |
| Physical activity<br>(MET-hour/week) | 19.80 [1.65, 48.25] | 25.55 [5.78, 54.79] | 22.85 [4.12, 51.10] | 19.32 [1.55, 47.55] | 12.38 [0.00, 37.77] | <0.001 |
| BMI (kg/m2) | 27.44 (4.78) | 22.86 (2.15) | 25.68 (2.07) | 28.10 (2.33) | 33.14 (4.46) | <0.001 |
| Waist(cm) | 90.00 [81.00, 99.00] | 75.00 [71.00, 79.00] | 86.00 [82.00, 90.00] | 94.00 [90.00, 98.00] | 105.00 [100.00, 112.00] | <0.001 |
| Sbp(mmHg) | 137.00 [125.00, 151.00] | 130.00 [119.00, 143.00] | 136.00 [125.00, 150.00] | 140.00 [128.00, 153.00] | 143.00 [131.00, 156.00] | <0.001 |
| Dbp(mmHg) | 82.00 [75.00, 89.00] | 77.00 [71.00, 84.00] | 81.00 [74.00, 88.00] | 83.00 [77.00, 90.00] | 85.00 [79.00, 92.00] | <0.001 |
| Log_CRP | 0.35 [-0.29, 1.04] | -0.24 [-0.73, 0.39] | 0.17 [-0.37, 0.78] | 0.48 [-0.07, 1.08] | 0.95 [0.36, 1.57] | <0.001 |
| TC (mmol/L) | 5.65 [4.91, 6.42] | 5.60 [4.94, 6.31] | 5.73 [5.02, 6.48] | 5.73 [4.96, 6.51] | 5.52 [4.69, 6.36] | <0.001 |
| LDL(mmol/L) | 1.75 [1.47, 2.06] | 1.70 [1.44, 1.98] | 1.79 [1.51, 2.09] | 1.80 [1.51, 2.12] | 1.73 [1.42, 2.06] | <0.001 |
| HDL(mmol/L) | 1.40 [1.17, 1.67] | 1.65 [1.41, 1.91] | 1.43 [1.22, 1.68] | 1.32 [1.12, 1.55] | 1.23 [1.05, 1.44] | <0.001 |
| HbA1c | 35.20 [32.70, 37.90] | 34.20 [31.90, 36.40] | 34.70 [32.40, 37.10] | 35.40 [33.00, 38.00] | 36.90 [34.10, 40.30] | <0.001 |
| Hypertension | 215803 (55.8) | 34639 (35.8) | 49042 (50.7) | 60104 (62.1) | 72018 (74.5) | <0.001 |
| Diabetes | 23474 (6.1) | 1804 (1.9) | 2932 (3.0) | 5345 (5.5) | 13393 (13.8) | <0.001 |
| Hyperlipidemia | 143898 (37.2) | 30743 (31.8) | 36885 (38.1) | 38429 (39.7) | 37841 (39.1) | <0.001 |
| Valve heart disease | 1340 (0.3) | 367 (0.4) | 340 (0.4) | 305 (0.3) | 328 (0.3) | 0.112 |
| Cardiovascular disease | 16639 (4.3) | 1932 (2.0) | 3218 (3.3) | 4508 (4.7) | 6981 (7.2) | <0.001 |
| Oral problems | 81441 (21.1) | 19019 (19.7) | 19310 (20.0) | 20197 (20.9) | 22915 (23.7) | <0.001 |
| Antihypertensive medication | 35392 (9.1) | 5461 (5.6) | 6961 (7.2) | 8209 (8.5) | 14761 (15.3) | <0.001 |
| Insulin | 1607 (0.4) | 304 (0.3) | 226 (0.2) | 233 (0.2) | 844 (0.9) | <0.001 |
| Cholesterol-lowering medication | 25562 (6.6) | 3764 (3.9) | 5100 (5.3) | 6142 (6.4) | 10556 (10.9) | <0.001 |

**Supplementary Fig. 1.**
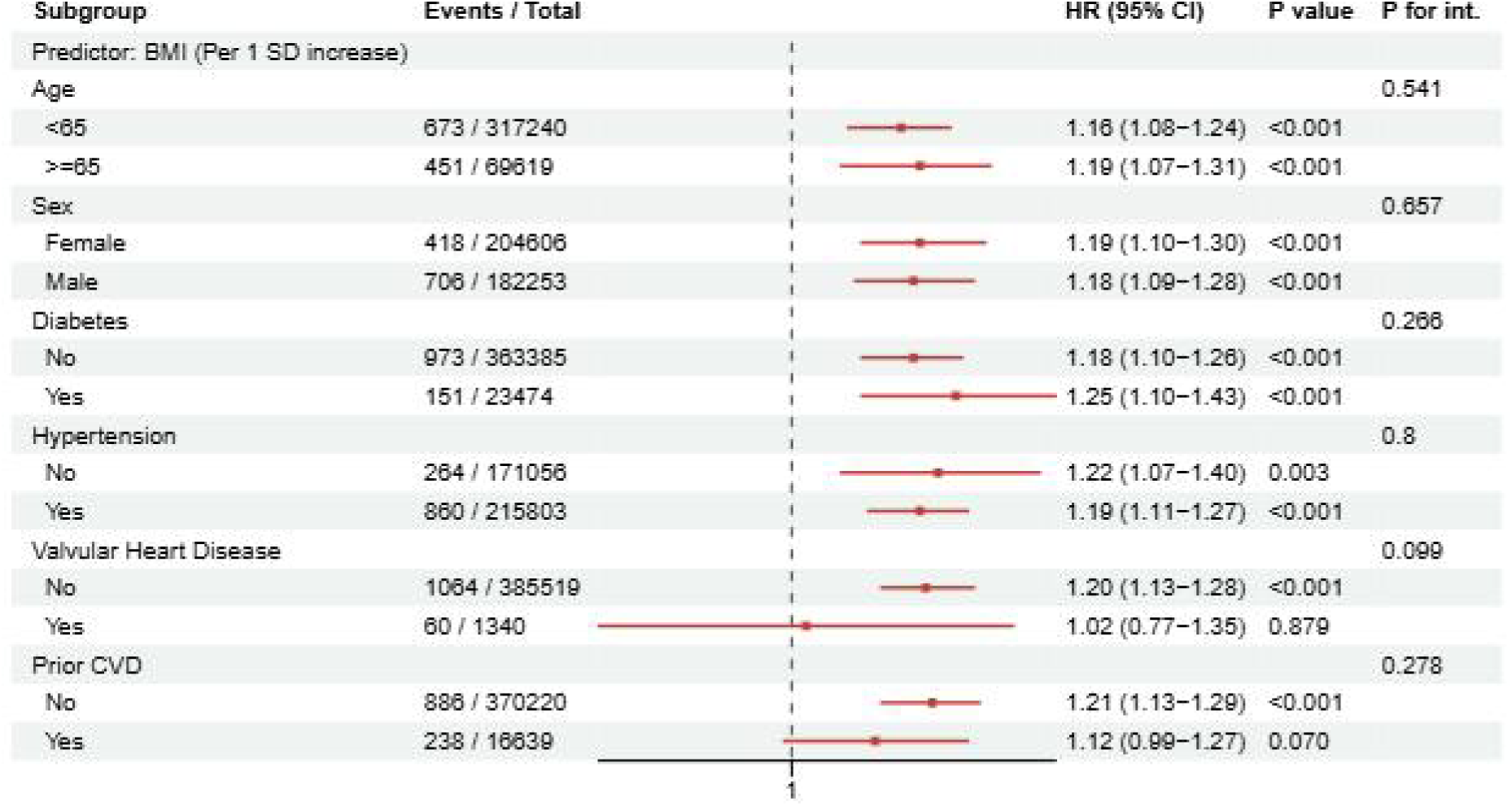
Subgroup and interaction analyses of the association between BMI and the risk of incident infective endocarditis. Adjusted for age, sex, race, education, Townsend deprivation index, smoking, drinking, physical activity, hypertension, diabetes, hypercholesterolemia, cardiovascular disease, valvular heart disease, oral problems. BMI, body mass index.

**Supplementary Fig. 2.**
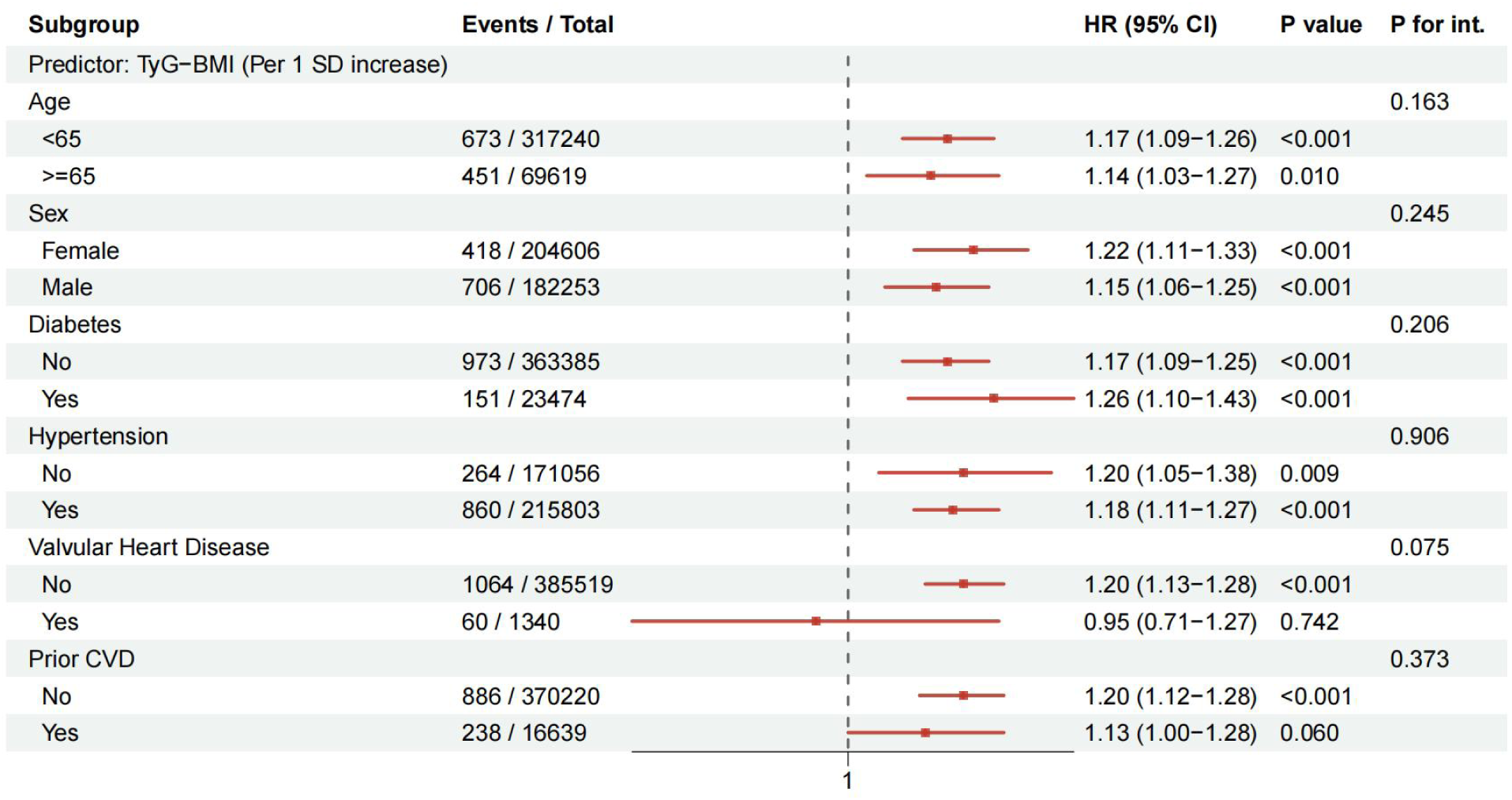
Subgroup and interaction analyses of the association between TyG-BMI and the risk of incident infective endocarditis. Adjusted for age, sex, race, education, Townsend deprivation index, smoking, drinking, physical activity, hypertension, diabetes, hypercholesterolemia, cardiovascular disease, valvular heart disease, oral problems. BMI, body mass index.

**Supplementary Fig. 3.**
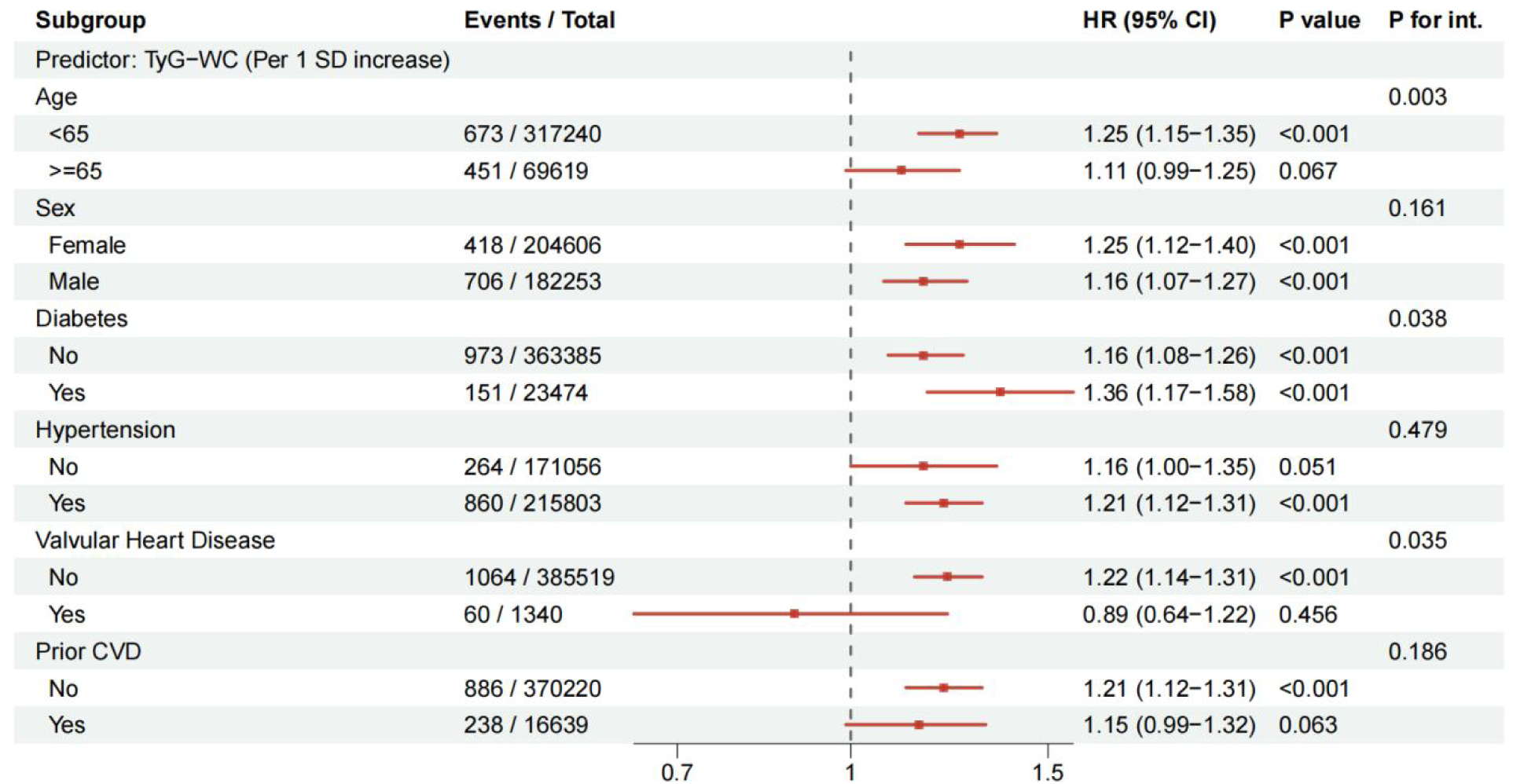
Subgroup and interaction analyses of the association between TyG-WC and the risk of incident infective endocarditis. Adjusted for age, sex, race, education, Townsend deprivation index, smoking, drinking, physical activity, hypertension, diabetes, hypercholesterolemia, cardiovascular disease, valvular heart disease, oral problems. BMI, body mass index.

**Supplementary Fig. 4.**
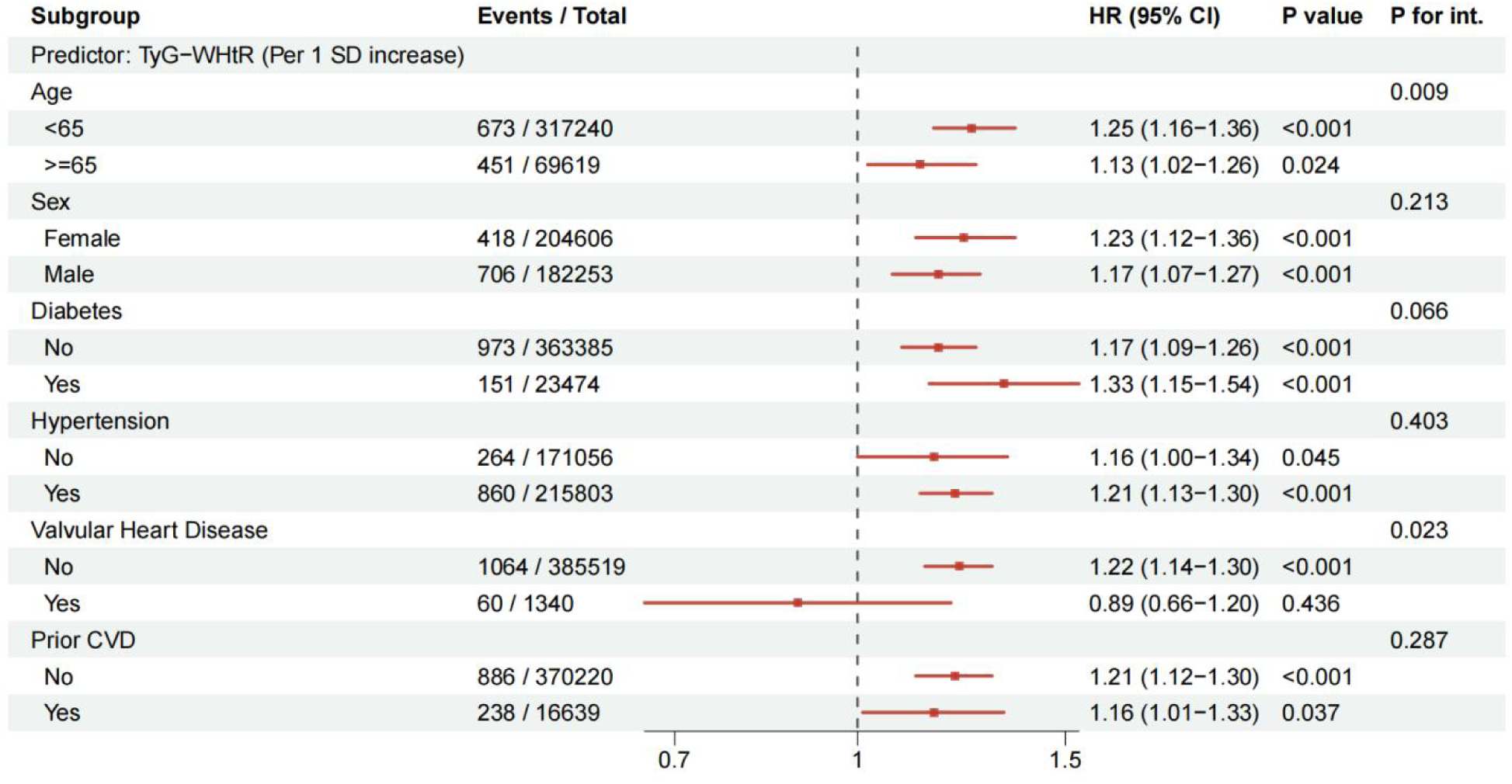
Subgroup and interaction analyses of the association between TyG-WHtR and the risk of incident infective endocarditis. Adjusted for age, sex, race, education, Townsend deprivation index, smoking, drinking, physical activity, hypertension, diabetes, hypercholesterolemia, cardiovascular disease, valvular heart disease, oral problems. BMI, body mass index.

**Supplementary Fig. 5.**
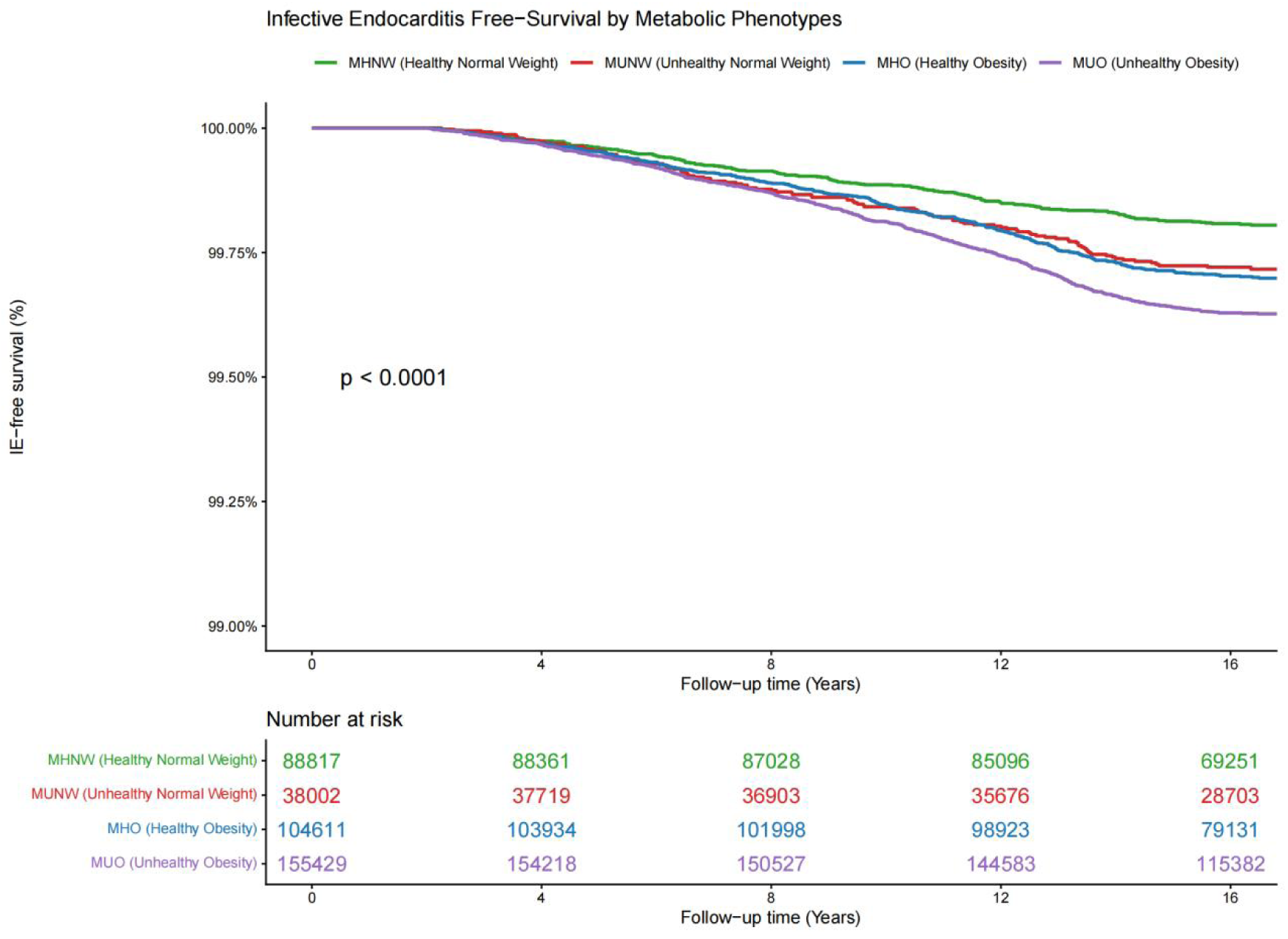
Kaplan-Meier survival curves for incident infective endocarditis stratified by metabolic obesity phenotypes. IE, infective endocarditis; MHO, metabolically healthy obesity; MHNW, metabolically healthy normal weight; MUO, metabolically unhealthy obesity.

**Supplementary Fig. 6.**
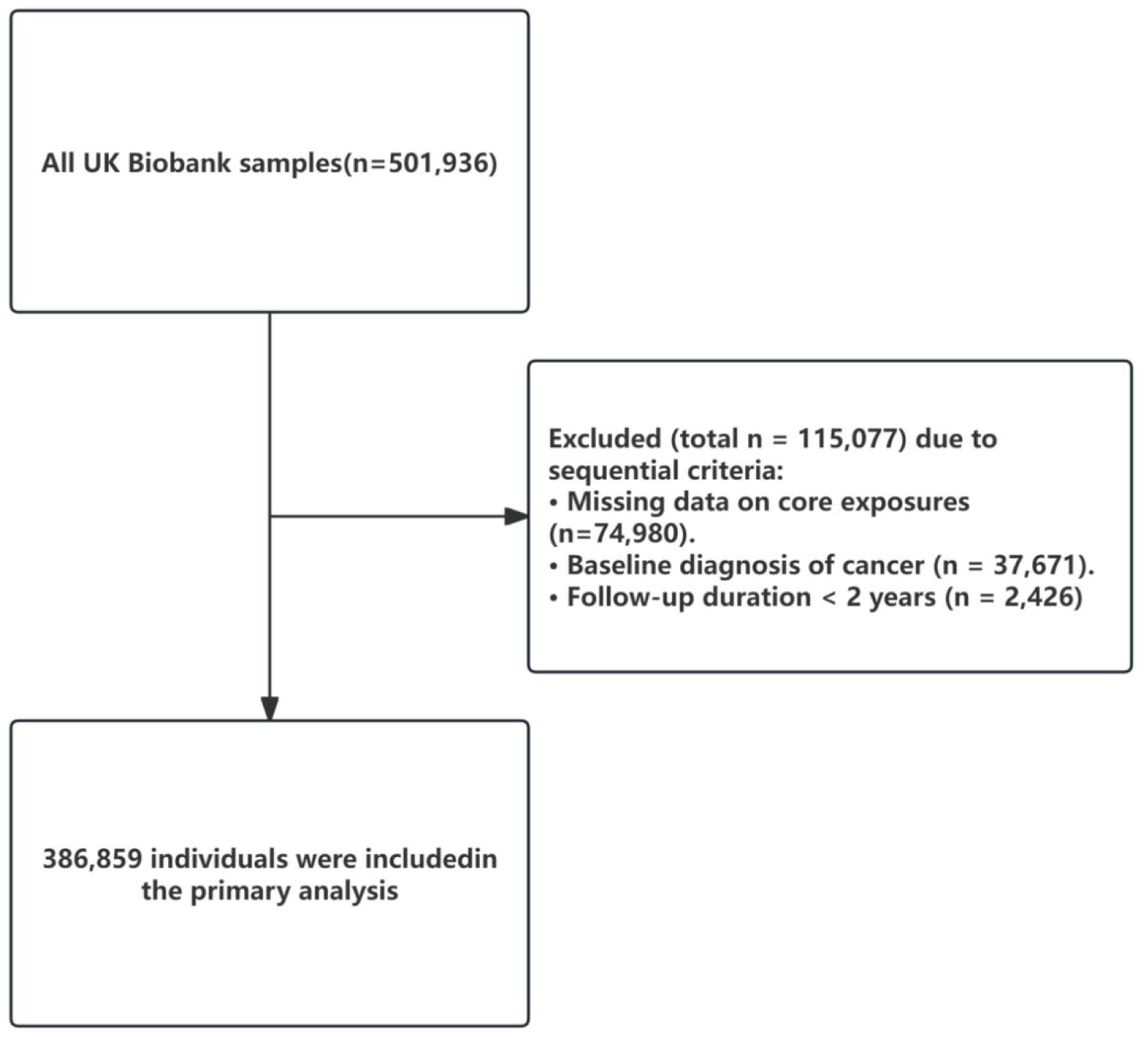
Flowchart of participant selection.

